# The MultiMusic multidomain intervention including choral practice in community-dwelling older people: A study protocol for Brain-Derived Neurotrophic Factor (BDNF) salivary biomarkers, audiometric and neuropsychological parameters

**DOI:** 10.1101/2024.11.29.24318152

**Authors:** M. Lippolis, R. Lenti, P. Mantuano, A. De Luca, A. Pantaleo, N. Quaranta, A. Pilotto, V. Solfrizzi, P. Vuust, E. Brattico

## Abstract

Brain-Derived Neurotrophic Factor (BDNF) plays a crucial role in neuroplasticity, supporting brain health along life and even during aging. This longitudinal study investigates the impact of a 9-month multidomain intervention, including choral practice, on BDNF levels, audiometric and neuropsychological outcomes, in older adults, assessed also for their multidimensional frailty, based on their physical, cognitive and nutritional conditions. BDNF levels, along with geriatric, neuropsychological, audiometric and neurophysiological measures, are taken, via saliva samples, both pre- and post-intervention. For BDNF longitudinal analyses, up to 80 elderly will be randomly assigned to an experimental group participating in a 9 month long multidomain program of activities including choir, physical, intellectual and manual activities or to an active control group participating in physical, intellectual or manual non-musical activities for 9 months or to a passive control group. Participation in daily activities for all groups will be monitored via diaries. Our primary goal is to investigate whether engaging in physical, cognitive and social multidomain activities can enhance neuroplasticity as measured by BDNF levels, to contrast aspects of multidimensional frailty (as assessed by the multidimensional prognostic score, MPI) in the aging population. Secondly, we aim to relate changes in BDNF levels to the perceptual and cognitive domains and psychosocial well-being. Although previous studies point out the positive effects of musical training and active aging on brain health, the scarcity of longitudinal research on effects on BDNF for older individuals keeps the issue open. Moreover, we aim to test whether non-invasive, accessible saliva-based BDNF measurements, despite some reliability limitations, could offer indications of neuroprotection in aging.

## Background

### 1. An “enriched environment” for a successful aging

Aging is a stage of life characterized by psychological and physiological decline, either in healthy or pathological conditions. While ‘crystallized’ skills (i.e., abilities learned throughout life like autobiographical memories and vocabulary), tend to remain stable, a decline is observed in fluid abilities, like executive functions (Binder et al.,, 2016). In the brain, aging is reflected by typical morphologic and functional changes, such as brain matter shrinkage and loss of synaptic strength, that have been related to the aging process (Taki et al., 2011; Morrison & Baxter, 2012). Few studies suggest that those neuronal changes may be related to the decrease of brain-derived neurotrophic factor (BDNF), a neurotrophin plays a role in maintaining brain function levels over the course of life (e.g., Zeng et al., 2011; Driscoll et al., 2012; Mizoguchi et al., 2020).

Although irreversible, however, the decline process related to aging may be slowed down; acting on modifiable risk factors (e.g., sedentariness, obesity, poor social skills, little cognitive stimulation etc.) may help prevent neurodegenerative diseases (Nazam et al., 2021). Older people can, indeed, engage in various compensatory and adaptive processes to maintain cognitive function. This adaptive process, known as “scaffolding” (Park and Reuter-Lorenz, 2009; 2014), helps individuals cope with neural decline thanks to experience-dependent plasticity, which can be enhanced through cognitive training, continuous social or intellectual engagement, learning new skills, and regular physical exercise. This is possible as experience-dependent neuroplasticity, which is most evident during childhood development, has been demonstrated to persist into adulthood and during aging (Boyke et al., 2008; Lövdén et al., 2010; Zatorre et al., 2012).

A “successful aging” occurs when elderly experience a high physical, psychological, and social functioning in absence of major diseases (Rowe & Kahn, 1997). On the contrary, when vulnerable and at risk, they are considered as “frails” (Italian National Institute of Health, 2023). Multidimensional frailty (Pilotto et al., 2020) raises the risk of adverse health outcomes, i.e., falls, hospitalization, institutionalization etc. (Clegg et al., 2013). Multidimensional frailty is closely related to quality of aging, as they are influenced by common factors and have a mutual impact. In fact, both rely on the domains of health and daily living activities, physical and cognitive operation, social participation and engagement (Urtamo, Jyväkorpi & Strandberg, 2019; Veronese et al., 2022). Consequently, these multiple domains need to be targeted with a multifaceted approach to intervention.

Some authors addressed the concept of “enriched environment” as a strategy to foster neuroprotection in aging (e.g., Mora, 2013; McDonald et al., 2018; Miranda et al. 2019) with the aim promote active and healthy aging, for rehabilitation or to prevent neurodegenerative diseases. Such a notion stems from the first animal studies conducted during the last century, where experimental settings were designed to enhance cognitive, social and motor ability and to potentiate learning and memory (Hebb, 1947; Rosenzweig et al., 1962; Diamond et al., 1964). In a broader sense, an enriched environment for humans implies the set up of contexts and conditions that foster personal development, social, emotional and cognitive enhancement, and general well-being.

One type of intervention akin to an enriched environment is the multi-domain intervention, which can be used to support healthy aging and reverse multidimensional frailty (Schneider & Yvon, 2013; Murukesu et al., 2020). Multi-domain intervention is commonly defined as a therapeutic or preventive strategy involving two or more domains (e.g., diet, physical activity, cognitive training etc.) to address complex or multifactorial problems. Thus far, it has been found to be particularly effective for healthy elderly (Rosenberg et al., 2018; 2020), able to induce brain changes in individuals at risk of developing Alzheimer’s disease (Perus et al., 2022) and to prevent frailty (Huang et al., 2023). Furthermore, the FINGER study by Ngandu et al. (2015) showed that a multidomain intervention can improve and maintain cognitive functioning in at-risk elderly people from the general population.

It is broadly known that engaging in challenging cognitive, physical and social activities can be a trigger for neuroplasticity and prevent multidimensional frailty and cognitive decline (Draganski et al., 2004; Glass et al., 2006; Cotman, Berchtold & Christie, 2007; Gerstorf et al., 2016; Pickersgill et al., 2022). Hence, the simultaneous combination of such a variety of stimuli may be considered a medium to prevent neurodegenerative diseases, despite transfer studies on the topic still being few and inconsistent (Binder et al., 2016). Some evidence on the effects of an intervention combining different domains on BDNF levels is recently emerging. For example, an exploratory study by Moon et al., (2022) evaluated the impact of two different 24-week multidomain interventions and found an increase in both serum BDNF levels and global cortical thickness for the multidomain group. However, the relationship between multidomain intervention and BDNF levels needs to be explored much more in depth, especially when considering the effects of a multidomain intervention including music training.

### 2. Brain-derived neurotrophic factor: Main roles and clinical implications

BDNF is a protein belonging to the neurotrophin family: a group of closely related proteins responsible for the survival, development, and function of neurons in both central and peripheral nervous systems (Reichardt, 2006). Discovered in 1982 (Adachi et al., 2014), BDNF plays a central role in many physiological processes including the establishment, strengthening, and maintenance of synaptic plasticity throughout life. Although its level increases during post-natal development, BDNF remains highly expressed in the adult brain and is involved in the regulation of both excitatory and inhibitory synaptic transmission, mainly by modulating in activity-dependent plasticity processes such as learning, and memory (Webster et al., 2006; Karpova, 2014; Kojima et al., 2017; Edelmann & Leßmann, 2018). Recent research has highlighted that the role of BDNF is not limited to brain development and aging, but is also crucial in pathological conditions such as depression, anxiety and neurodegenerative diseases. This has led to a surge of interest in BDNF as pharmacological agent and/or target over the past few decades (Colucci-D’Amato et al., 2020; Gao et al., 2022).

BDNF is mainly synthesised in the central nervous system (CNS), notably in some regions including the hippocampus, frontal cortex, midbrain, amygdala, hypothalamus, striatum, pons and medulla oblongata (Palasz et al., 2020; Lipsky & Marini, 2007; Suri et al., 2013). Furthermore, it is synthesised in peripheral non-neuronal cells, including T- and B-lymphocytes, monocytes (Wang et al., 2021), vascular endothelial cells (Meuchel et al., 2011), smooth and skeletal muscle cells (Matthews et al.,2009), adipocytes (Chaldakov et al., 2009), lacrimal glands (Ghinelli et al.,2003), and in the salivary glands of rats and humans (Saruta et al., 2010; Saruta et al., 2012). BDNF is stored primarily in a bound state within platelets in the blood, liver, and spleen. (Walsh et al., 2020).

The BDNF gene is located on chromosome 11 and contains 9 promoters that can initiate transcription of 24 transcripts, each containing an alternative 5′ noncoding exon spliced to a 3′ coding exon that comprises the entire open reading frame for the BDNF protein (Palasz et al., 2020). Similarly to other proteins, the synthesis of BDNF is a sequential process involving the formation of multiple precursor isoforms. A range of factors, including hypoxia, stress, epileptic seizures and ischemia, have been observed to increase BDNF expression (Brigadski & Leßmann, 2020). The preproBDNF precursor (32-35 kDa) is synthesized and folded in the endoplasmic reticulum, then translocated to the Golgi apparatus where the proregulatory sequence is cleaved to form proBDNF (28-32 kDa). ProBDNF can be converted in the trans-Golgi network into mBDNF (13 kDa) by the subtilisin-kexin family of endoproteases, such as furin, or into intracellular vesicles by convertases (Foltran & Diaz, 2016). Extracellular cleavage of proBDNF is mediated by plasmin and matrix metalloproteinases 2 and 9 (MMP2 and MMP9) (Liu et al., 2005; Mizoguchi et al., 2011; Kowiański et al., 2018).

BDNF functions as a paracrine and autocrine factor at both pre-synaptic and post-synaptic target sites. The proBDNF and mBDNF isoforms exhibit activity and stimulate opposing effects through interaction with the p75 neurotrophin receptor (p75NTR) and the TrkB receptor, respectively. These interactions stimulate signalling pathways involved in brain development, synaptic plasticity and protection or regeneration after damage. The balance between proBDNF and mBDNF differs depending on the stage of brain development and the region of the brain. During developmental stages, there is a more significant presence of proBDNF, while mBDNF is predominant in adulthood (Colucci-D’Amato et al., 2020).

A multitude of factors, including age, gender, body mass index (BMI) and gene polymorphism, may exert an influence on the levels of BDNF in brain, peripheral tissues, and blood. This also applies to environmental and hormonal factors such as caloric restriction (Mattson et al., 2003), estrogen levels (Scharfman & Maclusky, 2005) and environmental enrichment (van Praag et al., 2000). The single nucleotide polymorphism rs6265 o G196A located in the BDNF gene promoter, is responsible for a change in the amino acid sequence at codon 66 (Val66Met), resulting in a deficit in BDNF release and a reduction in the efficiency of BDNF-TrkB signaling, which is responsible for memory and LTP (Egan et al., 2003; Hariri et al., 2003; Zhao et al., 2018).

Reduced levels of BDNF have been linked to brain disorders such as Alzheimer’s, depression, and schizophrenia, as well as to the physiological aging process, which highlight its potential role in these conditions (Durany et al., 2001; Shimizu et al., 2003; Karege et al., 2005; Peng et al., 2005; Palomino et al., 2006), although some aspects are still controversial (Laske et al., 2006; Komulainen et al., 2008; Angelucci et al., 2009; Nettiksimmons et al., 2014).Considering the evidence reviewed above, a number of external interventions might boost BDNF expression. As a result, such interventions may represent viable treatment options for cognitive impairments associated with low BDNF levels age-related.

### 3. Music Training: A Powerful Catalyst for Brain Health

After a few decades of research, music training, intended as active music making with instruments or voice, is now recognised as a putative non-pharmacological intervention for structurally and functionally inducing neuroplastic processes in childhood, adulthood and aging (Zatorre, Chen, & Penhune, 2007; Herholz & Zatorre, 2012; Olszewska et al., 2020), making it a useful tool for both prevention and rehabilitation (Hyde et al., 2009; Seinfeld et al., 2013; Bergman-Nutley et al., 2014; Bidelman et al., 2014; Seither Preisler et al., 2014; Schlaug, 2015; Degé et al., 2018; MacAulay et al., 2019; Bugos et al., 2022; Lu et al., 2022; Lippolis et al., 2023; Marie et al., 2023; Schneider et al., 2023). In younger adults, music training results in functional changes such as significant modifications in synaptic strength across cortical networks, affecting various stages of the sensorimotor and auditory pathways as well as higher-order cognition areas (Lappe et al., 2008; Schlaug et al., 2009; Zuk et al., 2014; for a meta-analysis of neuroimaging studies, cf. Criscuolo et al., 2022). For older adults, long-term musical experience throughout life has been shown to act as a protective factor, as it leads to have an enhanced brain volume and cognitive functions in later life (White-Schwoch et al., 2013; Bidelman & Alain, 2015; Sihvonen et al., 2017; Chaddock-Heyman et al., 2021).

Besides duration, intensity of music training (i.e., actual time spent in musical practice) was shown to play a role. A study using diffusion tensor imaging showed enhanced structural connectivity in regions connecting visual, auditory, and motor areas due to the amount of individual musical practice (Ellis et al., 2013). Other studies, both on children and adults, which used intensity to define the music training variable, showed brain changes and increased abilities for those having a higher amount of practice, such as intellectual and verbal abilities (Loui et al., 2019), working memory (Bergman-Nutley et al., 2014), motor and auditory skills (Jabusch et al., 2009; Schlaug et al., 2009; Schneider et al., 2023). In older adults, intensity of musical practice also leads to an increased processing speed and visuospatial ability (Okely et al., 2023), phonological verbal fluency and reaction time (Wang et al., 2023). In particular, to be highly engaged in choral practice produces improvements in cognitive flexibility, phonemic fluency and sense of social integration (Pentikäinen et al., 2021; 2023).

For elderly, not only instrumental and choral training are beneficial from a cognitive point of view, but also for psychosocial well-being (Johnson et al., 2013; Coulton et al., 2015; Bugos et al., 2016; Biasutti & Mangiacotti, 2019; Pentikäinen et al., 2021; 2023; Zanto et al., 2023). This happens as musical activities help connecting with others, boosting self-esteem and reducing feelings of isolation and loneliness (Varvarigou et al., 2012). Additionally, these activities influence hormone levels, such as cortisol, and affect the autonomic nervous system by decreasing stress-related activation (de Witte et al., 2020).

The positive long-term effects of reiterated music listening and physical activities on BDNF are already known thanks to studies conducted on animals and humans (Chikahisa et al., 2006; Soya et al., 2007; Erickson et al., 2011; Forti et al., 2015; Xing et al., 2016; Schega et al, 2016; Nair et al., 2021), although, for both the evidence on physical activity is much more extensive than on music listening (cf. Brattico et al., 2021). The long-term effects have been studied in neuronal assemblies of animal models: repetitive and intensive electric stimulation induces long-term potentiation (LTP) mediated by NMDA glutamate receptors, leading to an increase of BDNF levels and to strengthening neuronal circuits at the synaptic level. Based on this, it is conceivable that repeated musical training could have similar effects at the cellular level in humans (Brattico et al., 2021). Supportive evidence of this hypothesis comes from a study by Minutillo et al. (2020): higher BDNF plasma levels were found in musicians’ brains as compared with non-musicians, providing a neurochemical explanation to the increased brain plasticity found in musicians by many neuroimaging studies (cf. Criscuolo et al., 2022). The picture becomes even more complex when considering the BDNF gene, which regulates the baseline levels of BDNF in humans.

Bonetti et al., (2023) demonstrated that music-derived plasticity in the auditory cortex (as reflected in enhanced brain MMN responses) was obtained only in Val/Val musicians as compared to non-musicians, whereas Met musicians did not differ from non-musicians in their auditory-cortex responses. However, this initial evidence in humans of a relation between music-derived plasticity and BDNF levels is confined to observational cross-sectional studies. To date, no longitudinal studies investigating the effects of music training on changes in BDNF levels.

### 4. How do we measure BDNF? A glance back at literature

The potential of BDNF as a biomarker in the diagnosis and evaluation of treatment outcomes for various diseases, including those associated with the aging process, is currently under wide investigation. Therefore, the availability of kits that permit the accurate and reproducible assaying of BDNF is paramount. The concentration of BDNF varies throughout lifespan, with generally higher levels observed during childhood compared to adulthood. However, BDNF levels tend to be higher in early adulthood (Webster et al., 2006), with a gradual decrease observed over time. Lommatzch and colleagues (2005) stratified average plasma levels of BDNF according to age group. In young adults, approximately 100 pg/ml was found (n = 45; age range 20–33 years). In middle age, the average plasma BDNF level was approximately 80 pg/ml (n = 59, age 34-47 years). In the elderly, the average plasma BDNF level was approximately 50 pg/ml (n = 36, age 48-60 years). Furthermore, a study of 5,104 adults aged 65-97 years demonstrated an average serum BDNF level of 21 ng/ml (SD = 5.4 ng/ml), exhibiting a decline of approximately 0.1 ng/ml for each additional year of age (Shimada et al., 2014). BDNF can permeate the blood-brain barrier in both directions, thereby exhibiting a bidirectional correlation between peripheral and cerebral BDNF levels (Ikenouchi et al., 2023).

The quantification of BDNF in whole blood, plasma and serum can be performed with an enzyme immunoassay (ELISA). It is recommended that serum is used as the preferred sample, as BDNF is approximately 100-fold higher than in plasma. Furthermore, plasma concentration is influenced by the handling of blood samples due to the presence of platelets, which store BDNF and can secrete it. Indeed, during the process of blood collection, the stress (or the damage) generated by the needle may cause platelet degranulation with the consequent secretion of BDNF. Additionally, the time taken by the operator to separate the plasma, or changes in ambient temperature, may also induce platelet degranulation and a significant release of BDNF into the plasma fraction.

Additionally, the release of BDNF from platelets may be influenced by individual factors, such as age, disease, or drug treatment. Consequently, the quantification of BDNF from plasma is susceptible to variations in sample preparation procedures and is challenging to reproduce among different operators (Polacchini et al., 2015). The strict adherence to standard operating procedures is therefore required for reliable plasma determinations. Similarly, the measurements of BDNF in whole blood samples is challenging. In fact, although results comparable to those obtained in serum can be obtained, the preparation of blood samples involves a methodological step of cell lysis, which introduces an additional variable (Elfving et al., 2010). In general, the process of blood sampling in certain patient groups, such as children and the elderly, can be perceived as stressful and time-consuming. Therefore, the development of a less invasive procedure for the determination of BDNF concentration would be advantageous.

A potentially viable alternative is salivary BDNF. Saliva sampling offers several advantages. Firstly, it is a minimally invasive procedure that allows a reasonable amount of sample to be collected, also multiple times. Secondly, although it is preferable for the procedure to be rigorously performed by experienced personnel, it may be performed also independently, making it accessible to a wider range of patients, including children and elderly people (Pandit et al., 2024) In support of this approach, whole saliva is a complex mixture of secretions from salivary glands and blood components that pass through intercellular spaces by transcellular or paracellular diffusion into saliva. Consequently, many substances present in blood are also present in saliva, including epidermal growth factor, nerve growth factor and insulin growth factor (Jasim et al., 2018; Kagami et al., 2000). Recent studies have demonstrated the presence of BDNF in human saliva using immunoblotting and enzymatic digestion (Mandel et al., 2009; Mandel et al., 2011). However, while several commercial ELISA kits have been validated for the measurement of BDNF in plasma and serum, none have yet been validated for the analysis of salivary BDNF. Hence, a secondary scope of this protocol is to achieve a validation of salivary BDNF as extracted with ELISA.

### 5. Aims of the study

Aging is a delicate phase of life, with important physical, cognitive and functional changes. In Italy, there is a high prevalence of secondary aging, due to poor prevention and poor early intervention on modifiable risk factors (Guastafierro et al., 2020). However, with the lengthening of human life span, old age is now seen not only from the perspective of decline. It is instead considered a stage of life worthy of being kept active and meaningful. Therefore, an increasing number of programmes are being implemented for the psycho-physical well-being of people even in later life (Poscia et al., 2017; Barbabella et al., 2022).

With the present study, we aim to examine the effectiveness of a non-pharmacological and multi-domain intervention including participation in choir for musically naïve healthy elderly, while validating the methodological approach for extracting BDNF from saliva. Given the existing literature on the topic, our main objective is to test whether a reiterated, intensive participation in several motor, manual, intellectual and musical activities can enhance BDNF levels in older adults, along with neuropsychological and neurophysiological outcomes including risk of multidimensional frailty. Our idea is that continuity and assiduity in attending enriching contexts, from both cognitive, social and physical point of view, can promote neuroplasticity and contrast multidimensional frailty in aging, and that this is linked to perceptual, cognitive and socio-psychological well-being.

### 6. Research hypothesis

We intend to exhibit for the first time that a multi-domain intervention including choir can counteract cognitive and perceptual-auditory decline and frailty in aging, supported by BDNF-related neuroplastic processes. Specifically, we hypothesize to find significantly increased BDNF levels over time in the intervention group, hand in hand with cognitive and perceptual improvement (measured with performance tests and questionnaires) with respect to a control group carrying out only sporadic activities. In the case of the individuals scoring higher in multidimentional frailty, we identify an improvement in the slowing of cognitive and perceptual decline over the course of time.

## Methods

The trial is registered in Clinicaltrials.gov public registry under the ID NCT06767410. We adopted rigorous research protocols to ensure the validity of the results. For the sake of brevity and clarity, we summarized in **Table 1** the most relevant information and details inspired by WHO guidelines (World Health Assembly, 2022). The study, including the BDNF sample collection procedures, has already been approved by the institutional review board of the department of Education, Psychology and Communication of the University of Bari (reference number: ET-23-27). A unique ID was created for each participant to ensure the anonymity of data. Moreover, due to the wide range of measurements envisaged for a comprehensive evaluation of participants, an interdisciplinary collaboration was set up between the Department of Education, Psychology, Communication and the Department of Pharmacology at the University of Bari, as well as with the Otolaryngology and Geriatrics Units of the Policlinic consortium hospital of the University of Bari.

**Table 1.**
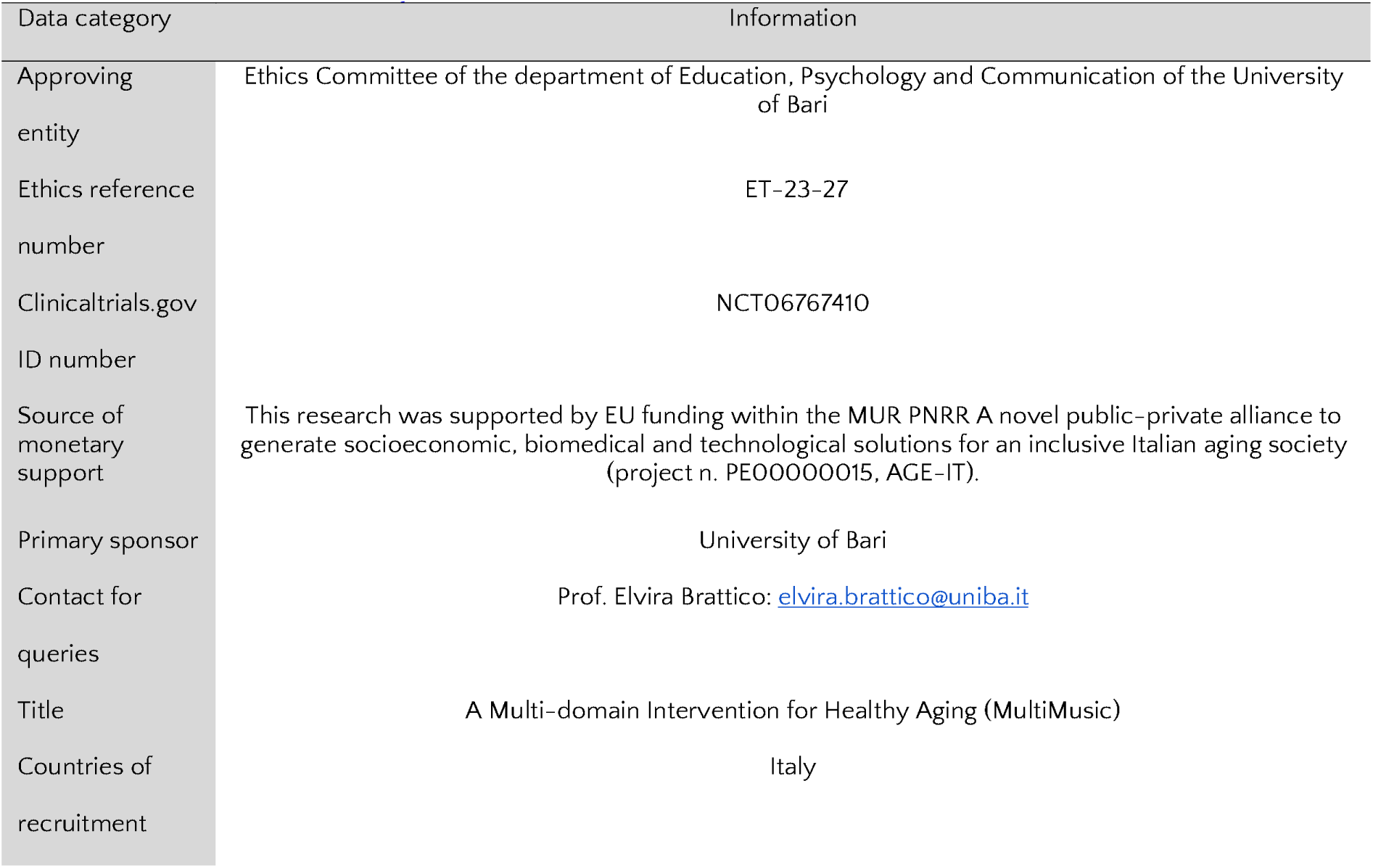
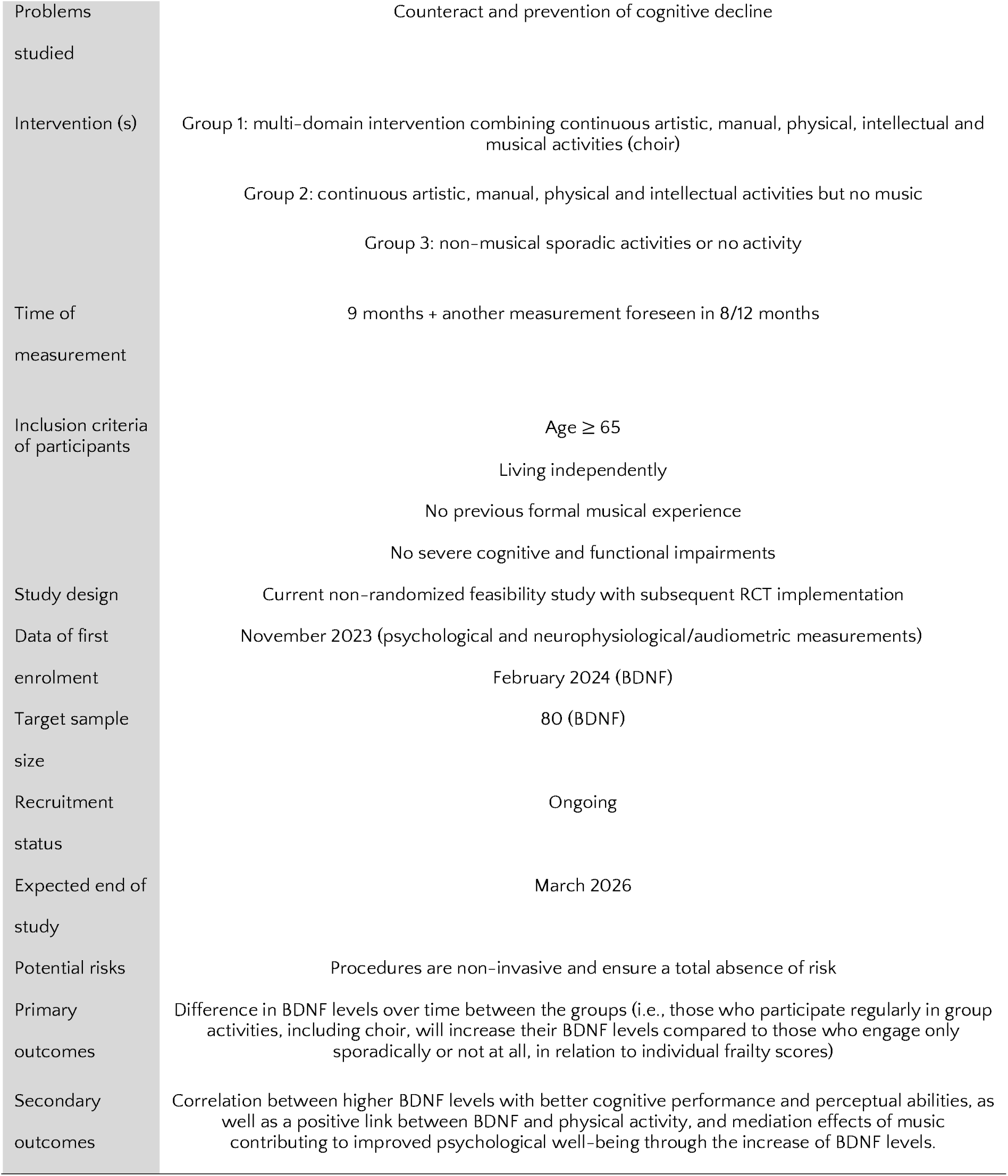
Information about the study.

|  |  |
| --- | --- |
| Problems studied | Counteract and prevention of cognitive decline |
| Intervention (s) | Group 1: multi-domain intervention combining continuous artistic, manual, physical, intellectual and musical activities (choir)<br>Group 2: continuous artistic, manual, physical and intellectual activities but no music<br>Group 3: non-musical sporadic activities or no activity |
| Time of measurement | 9 months + another measurement foreseen in 8/12 months |
| Inclusion criteria of participants | Age $\geq 65$<br>Living independently<br>No previous formal musical experience<br>No severe cognitive and functional impairments |
| Study design | Current non-randomized feasibility study with subsequent RCT implementation |
| Data of first enrolment | November 2023 (psychological and neurophysiological/audiometric measurements)<br>February 2024 (BDNF) |
| Target sample size | 80 (BDNF) |
| Recruitment status | Ongoing |
| Expected end of study | March 2026 |
| Potential risks | Procedures are non-invasive and ensure a total absence of risk |
| Primary outcomes | Difference in BDNF levels over time between the groups (i.e., those who participate regularly in group activities, including choir, will increase their BDNF levels compared to those who engage only sporadically or not at all, in relation to individual frailty scores) |
| Secondary outcomes | Correlation between higher BDNF levels with better cognitive performance and perceptual abilities, as well as a positive link between BDNF and physical activity, and mediation effects of music contributing to improved psychological well-being through the increase of BDNF levels. |

### 1. Design and timeline

The present study is taking place in Bari, Southern Italy. A pilot non-randomized feasibility pilot study of the multi-domain intervention on a small sample is currently underway, with a randomized controlled trial (RCT) planned for subsequent implementation which will maintain the same features apart from random assignment of participants to the three groups.

The RCT study employs a longitudinal research design on the effects of a multi-domain intervention and includes an active group and a control group. The first group carries out a repeated, regular series of various musical, physical, intellectual and manual activities, while the second one engages only in sporadic activities and the third group does not undertake any activity. Each participant is tested 9 months after the first measurement. In addition, another measurement time is planned after a further 8-12 months. The study design is summarized in **Figure 1**.

**Figure 1.**
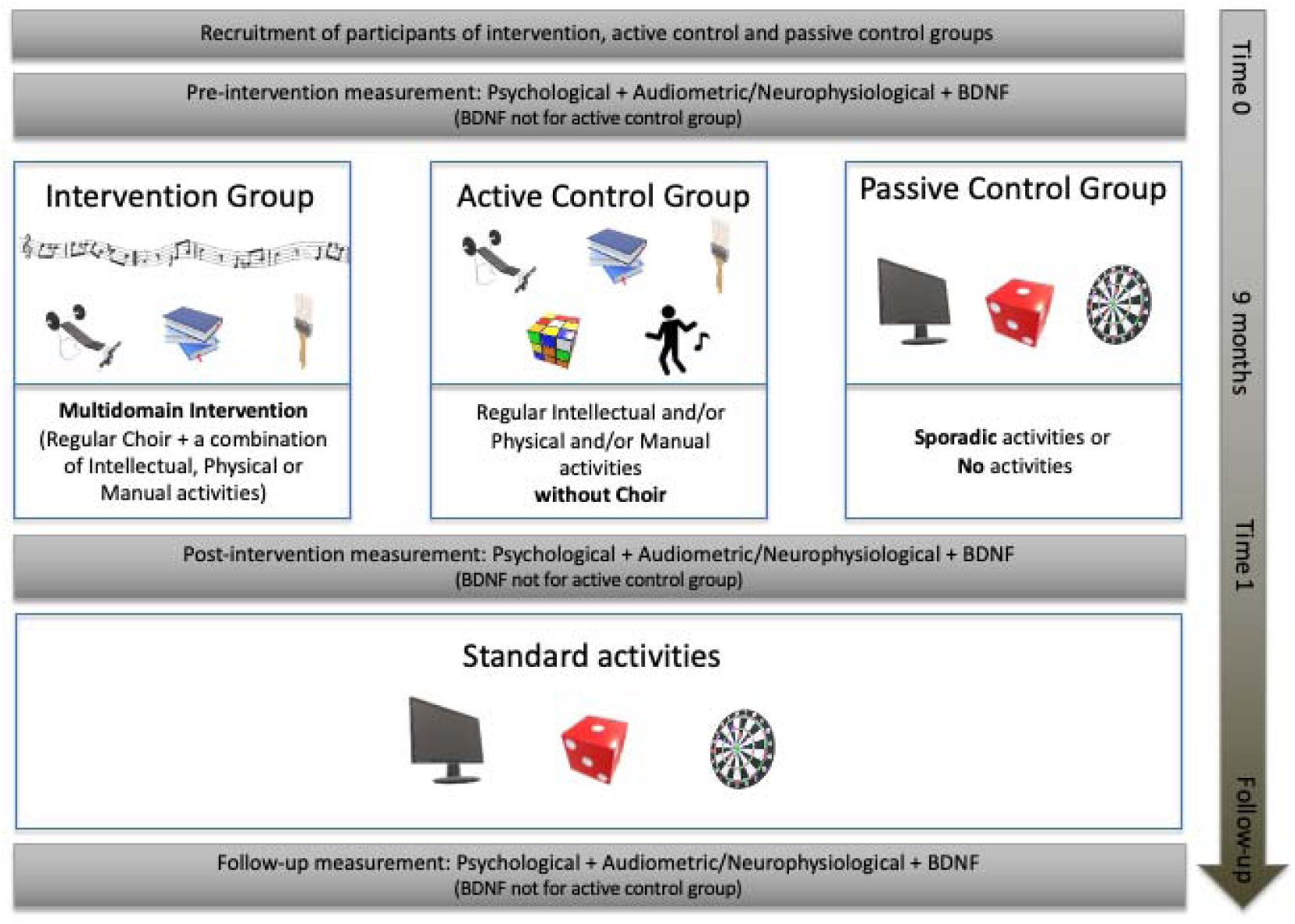
Study design

For the pilot feasibility study, participants from the two active groups freely choose which activities they want to attend. They have been being recruited since November 2023 through some district service centers for elderly people located both in the metropolitan and suburban areas. Also for the RCT, we will reach elderly participants via district service centers but we will randomly assign them to the three groups.

### 2. Participants, inclusion/exclusion criteria

Participants are included in the study if they are 65 or older, live independently, have no continuing and extensive musical experience and do not have severe cognitive or functional deficits. The intervention group attends multiple activities carried out in synergy and in dedicated centers, including choral training. The passive control group participates only in sporadic (non-musical) activities or no activities. The study also includes an active control group made of participants involved in several activities but not in choir.

This latter group, however, is not included in the sampling for the analysis of BDNF levels as the potential variability in salivary BDNF concentrations across different active groups could obscure the specific effects of the experimental intervention. Indeed, it has already been documented that detecting BDNF in saliva presents significant challenges due to its relatively low concentration and the influence of various external factors, making it difficult to draw clear conclusions from salivary measurements alone (Vrijen et al., 2017).

### 3. Intervention

The choral technique and singing class has been running for two hours a week since summer 2023. At the beginning of each meeting, the teacher starts with body percussion exercises carried out playfully and designed to strengthen body awareness, memory, motor coordination and concentration. Subsequently, respiration exercises and explanations of the functioning of organs impaired in the phonation process are proposed, in order to raise awareness on the use of vocal and respiratory apparatus. This is followed by progressive vocalizations, adapted to the textures of the various vocal sections (basses, tenors, altos, sopranos and treble voices) and, finally, the repertoire is studied, starting with participants’ cultural background, i.e. music from popular culture, then gradually enriched it with more complex, multi-voice and more articulate pieces. Songs are also performed with the participation of soloists and also in the form of duets and trios, in dialogue with the choir, to stimulate creativity and improvisation. Simple musical and rhythmic reading exercises are also provided, being participants totally naïve in musical instruction.

As here we deal with a multidomain intervention, participants are also engaged in various other non-musical activities. **Table 2a** contains a list of all these activities, also specifying frequency and intensity of each of them. **Table 2b** contains a summary of how programmes are currently distributed among participants and how they are combined, as an example for the future RCT. Moreover, diaries of the activities followed by each participant will be kept for accurately monitoring the intensity of the multi-domain intervention. Activities are grouped into the following macro-categories:

- Physical activities: this category includes gymnastics, group dancing, sports (e.g., swimming), postural exercise (pilates or body harmony).
- Musical activities: choir and body percussion.
- Manual activities: horticulture, handicraft, tailoring, cooking.
- Arts/Intellectual activities: theater, mental gymnastics, reading, learning programs (such as history etc.).

**Table 2a.**
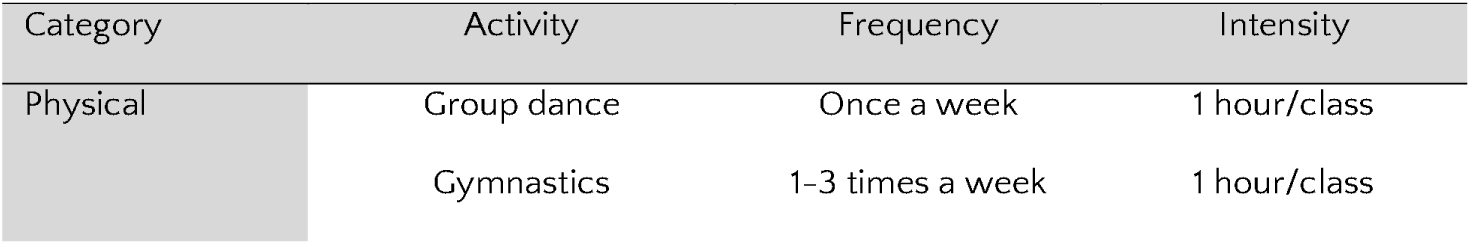
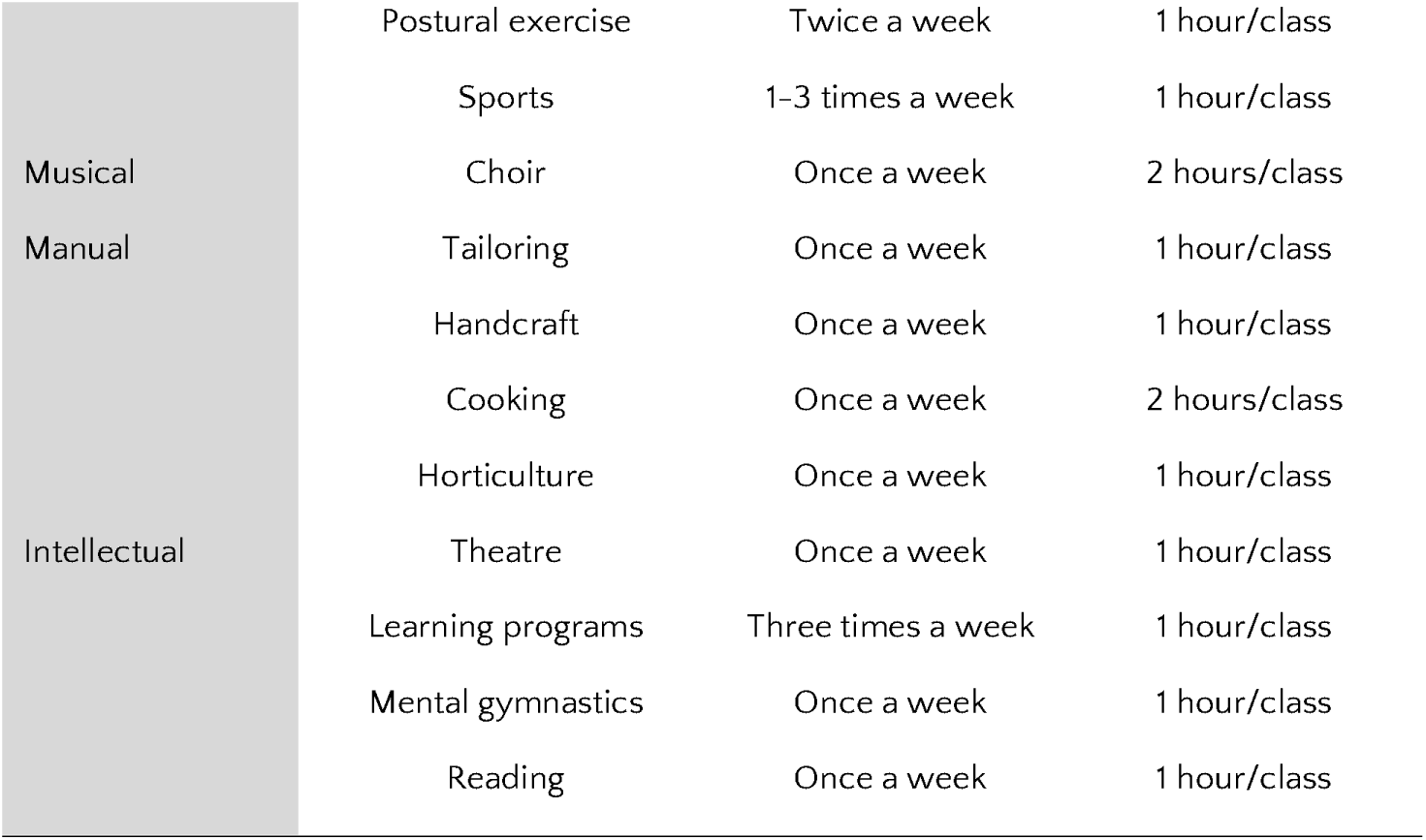
List of activities carried out by the participants.

**Table 2b.**
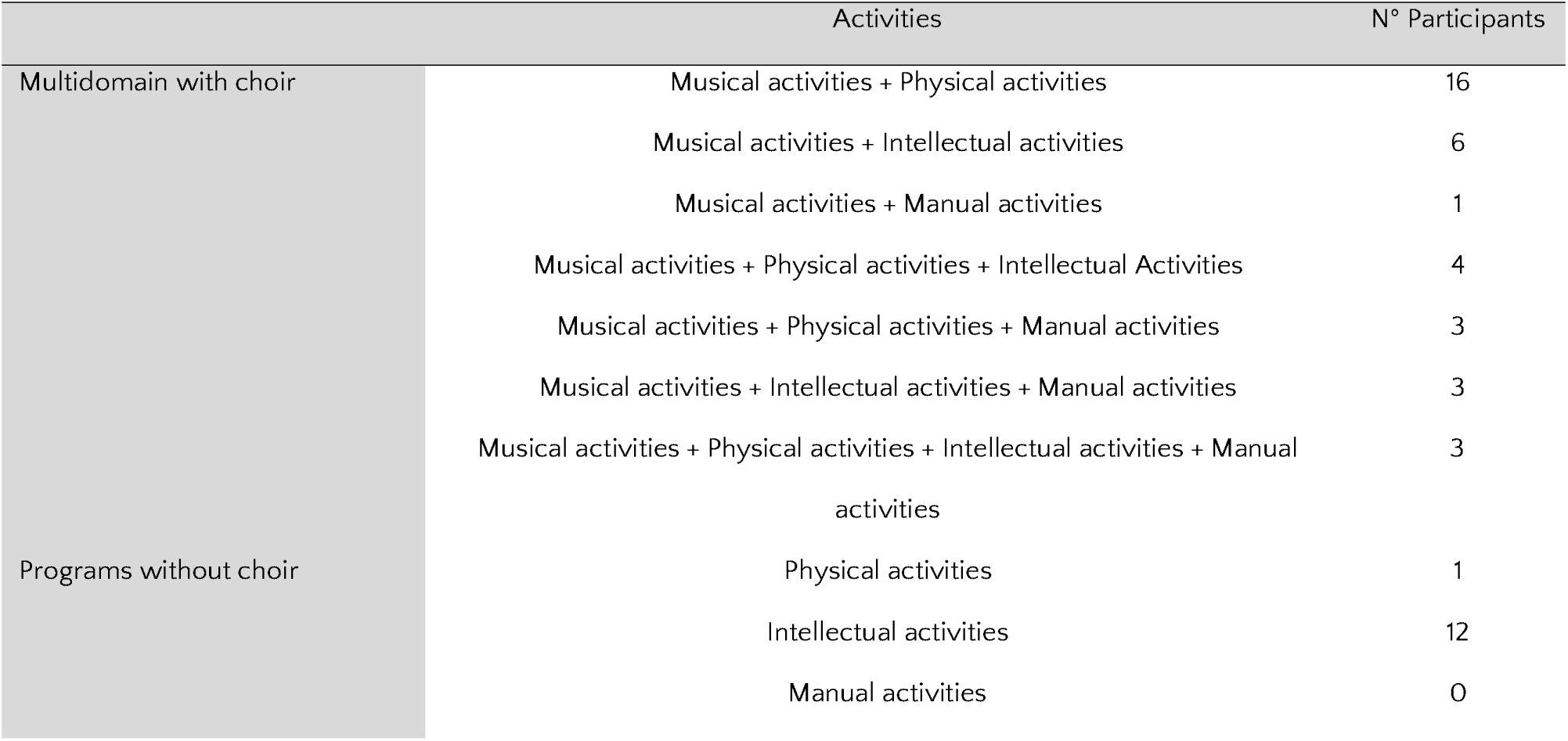
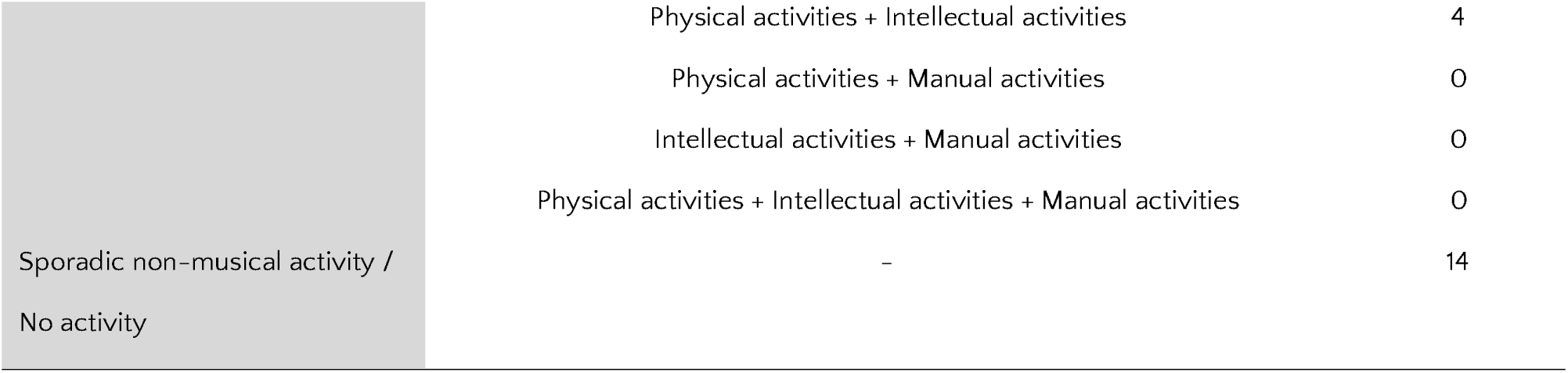
Distribution of activities among participants.

### 4. Procedures and measurements

#### a) Saliva collection

Prior to sample collection, participants sign informed consent forms to perform sample collection. They were instructed to abstain from eating, drinking, or brushing their teeth for two hours before the collection, as well as from consuming alcoholic beverages for 24 hours; this, in order to avoid any alteration or contamination of the samples. Right before saliva collection, each participant rinsed their mouth with distilled water in order to remove debris and moisturize the oral mucosa. Following a period of ten minutes, samples were collected via passive autonomous salivation. This approach was adopted in order to prevent the potential for non-specific binding of proteins present in the samples by cotton within the collection devices (ex. Salivettes). This phenomenon can result in a reduction in the concentration of these proteins including BDNF (Mandel et al., 2011). Approximately 1 mL of saliva was collected from each participant in a pre-cooled Eppendorf tube; a protease inhibitor cocktail (v/v 1:500 PMSF, Sigma Aldrich, St. Louis, MO, USA) was added right after collection.

#### b) Behavioral measurements

Besides BDNF sample collection, participants underwent a range of behavioral measurements (summarized in **Table 4**) to assess various domains. Cognitive functions, including visual and sustained attention, task switching, visuoconstructive skills, naming, memory, working memory, calculation, language, fluency, abstraction, and orientation, were evaluated using the Montreal Cognitive Assessment (MoCA) by Nasreddine et al. (2005). Each section includes specific tasks, such as word recall, solving logical problems, drawing figures, and naming objects.

**Table 3.**
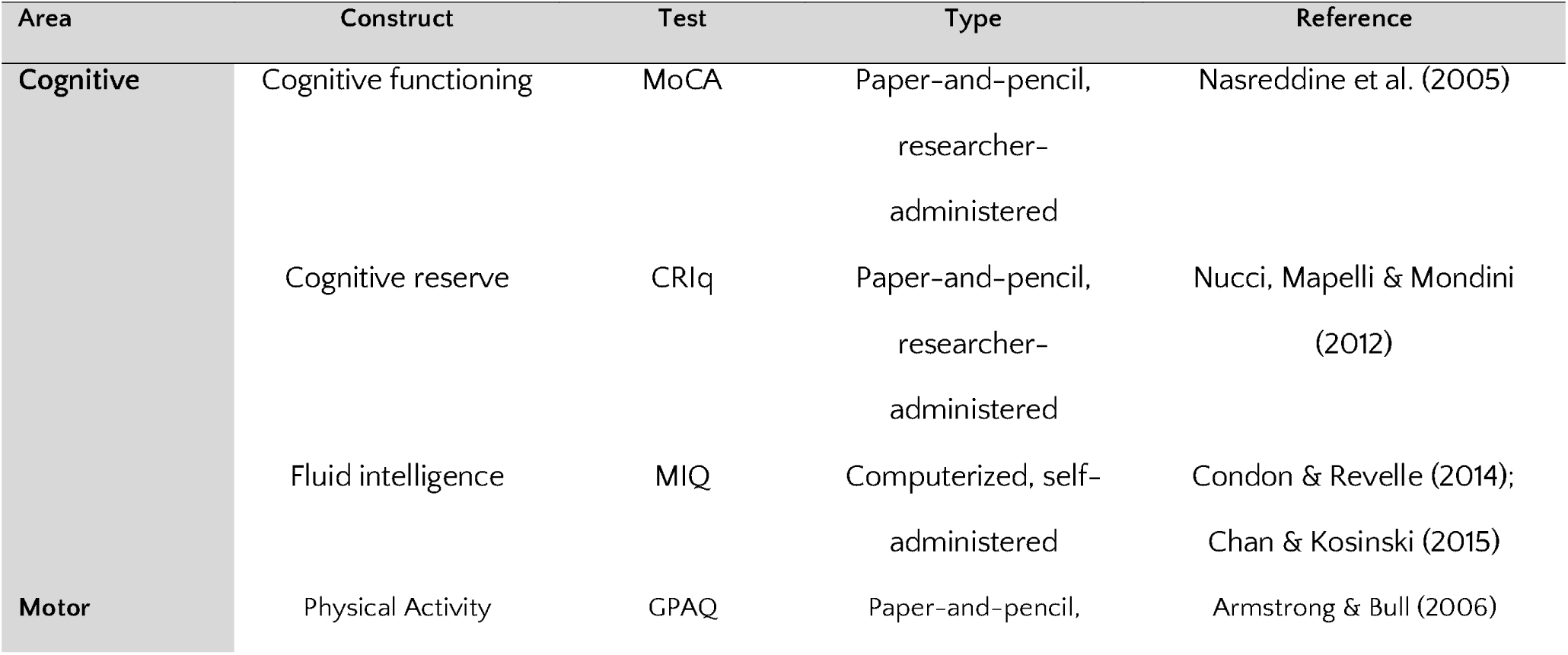
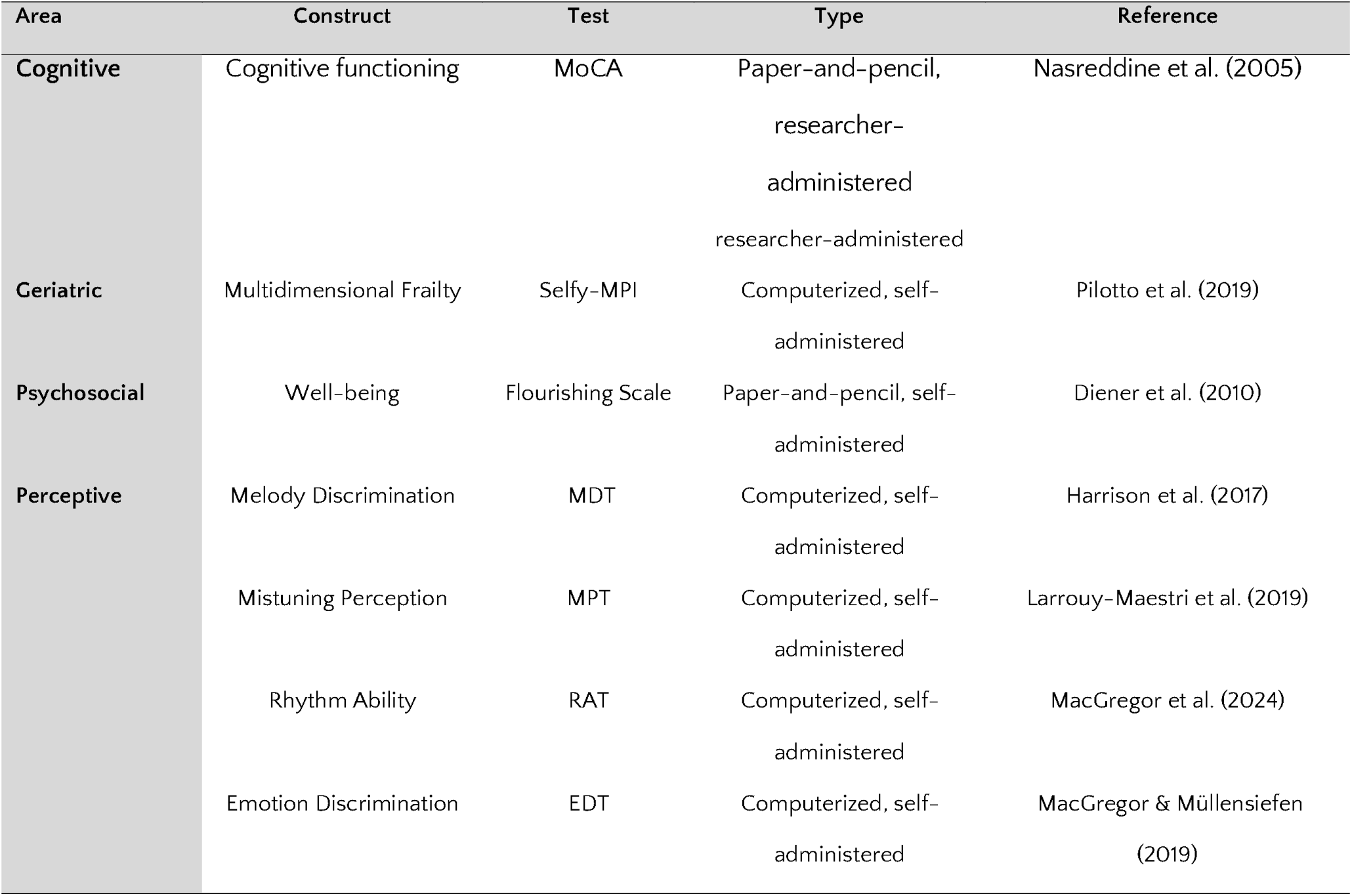
Behavioral assessment. List of the standardized tests and questionnaires used for assessing psychological functions of the elderly participants.

**Table 4.** A Priori Power Analysis. Power size analysis conducted with Jamovi.

| N<sub>1</sub> | N<sub>2</sub> | User Defined |  |  |
| --- | --- | --- | --- | --- |
| | | Effect Size | Power | $\alpha$ |
| 26 | 26 | 0.80 | 0.80 | 0.05 |

Cognitive reserve was measured with the Cognitive Reserve Index Questionnaire (CRIq) developed by Nucci, Mapelli & Mondini (2012), based on factors such as education level, professional career, and engagement in intellectual and social activities. Physical activity was monitored using the Global Physical Activity Questionnaire (GPAQ) by Armstrong & Bull (2006), which measures the frequency and duration of physical activities in different areas of life, including work, daily walking and recreational activities.

New computerized adaptive tests were used to assess both fluid intelligence (Matrix Reasoning, Condon & Revelle 2014; Chan & Kosinski, 2015) and musical abilities. Specifically, the perceptive tests used were created by the LongGold group: the Melody Discrimination Test (MDT; Harrison et al., 2017), the Rhythm Ability Test (RAT; MacGregor et al., 2024), the Mistuning Perception Test (MPT; Larrouy-Maestri et al., 2019) and the Emotion Discrimination Test (EDT; MacGregor & Müllensiefen, 2019).

In the Matrix Reasoning task (MIQ) participants identify the missing shape in a 3×3 array of geometric shapes, similarly to Raven’s Progressive Matrices. The Melody Discrimination Test (MDT) is a three-alternative forced-choice test where participants identify the "odd" melody out of three, based on similarity judgments. In the Rhythm Ability Test (RAT), participants select the image that matches the rhythm they hear, with increasing difficulty and sounds of varying pitches. In the Mistuning Perception Test (MPT): participants distinguish between in-tune and out-of-tune musical excerpts, using a two-alternative forced choice format. In the Emotion Discrimination Test (EDT), participants identify the emotion expressed in a melody (anger, happiness, sadness, or tenderness) by comparing two musical fragments, again in a two-alternative forced-choice format.

For the sake of accuracy in results, attendance of any extra-musical activity and/or individual intensity of choral practice were also assessed. Socio-psychological prosperity was measured using the Flourishing Scale by Diener et al. (2010), while multidimensional frailty was assessed with the Selfy-MPI (Pilotto et al., 2019).

The Selfy-MPI is a self-administered instrument here used in its digital version via the Portable-MPI app (downloadable from the following webpage: https://multiplat-age.it/index.php/it/strumenti) and is based on several dimensions:

- Functional status and independence, measured respectively by ADLs (Activities of Daily Living) and IADLs (Instrumental Activities of Daily Living), the first one measuring the ability to perform basic daily activities, such as dressing and eating, while the second one assesses more complex activities that require a certain level of independence, such as shopping and managing finances.
- Mobility, assessed through the MOB, a series of self-assessment questions concerning the ability to move independently.
- Cognitive condition, measure with brief test assessing temporal and spatial orientation, short-term memory ability, calculation, language and attention.
- Nutritional status, assessed with MNA (Mini Nutritional Assessment), including information on anthropometric measures (body mass index and weight loss), neuropsychological problems, recent psychological stress, mobility, and decline in food intake.
- Comorbidity, i.e., assessment of the number and severity of chronic diseases using the CIRS (Cumulative Illness Rating Scale).
- Medication use, through the Anatomical Therapeutic Chemical Classification code system (ATC classification), where the participant has to state the number of drugs taken regularly to detect the presence of poly pharmacotherapy.
- Living situation, through questions asking if the participant is living alone or with family members, at home or in RSA.

After the completion of all the sessions an automatic score is computed. Values between 0 and 0.33 indicate low prognostic frail risk; values between 0.34 and 0.66 indicate moderate prognostic frail risk; values between 0.67 and 1.00 indicate severe prognostic frail risk.

The psychological measurements are taking place in the presence of researchers and qualified psychologists, in those leisure centers collaborating in the research with a duration of about 1.30 hours per participant. Participants are tested individually or in small groups, often in pairs, alternating between computerized and paper-based tests and allowing for a more efficient use of time and resources. Additionally, participants were given careful instructions before beginning the tasks, as many were unfamiliar with technology, ensuring they were able to complete the computerized procedure independently.

#### c) Audiometric measurements

Tone and speech audiometric measurements are also taken (i.e.,liminal tonal audiometric test and speech audiometry), including a matrix sentence test to assess patients’ speech comprehension under noise conditions, and assessments of ABR and P300 auditory cortical potentials. The audiometric and neurophysiological measurements, as well as the BDNF sample collection, take place at the Otolaryngology Unit, lasting approximately 30 minutes per participant.

Liminal tonal audiometric test is used to determine the hearing threshold, i.e. the minimum acoustic intensity perceivable by the test subject at the different frequencies of the tonal field. The speech audiometry assesses and establishes verbal intelligibility, i.e. the recognition of words as a sound stimulus with meaning by sending the vocal material (phonemes, logotomes, words, sentences) to the patient in a silent booth.

The Matrix Sentence test is an adaptive test assessing the patient’s verbal comprehension under noise conditions, simulating the difficult listening conditions one may find oneself in in everyday life. The patient is asked to repeat 5-word sentences that are presented to him/her together with a disturbing noise (called ‘competition noise’). Likewise the perceptive computerized tests here used, this task is adaptive as that after each response the software automatically makes the next sentence ‘easier’ or ‘harder’ to understand. In this way, the test tends towards a precise volume of words and a precise volume of noise for which the patient will understand approximately 50% of the words presented. The result is a value known as the Signal Reception Threshold (SRT), which reflects the difference between the word volume and the signal volume at which the test reaches convergence.

The auditory evoked potentials of the brainstem are evoked by means of an acoustic stimulus (click) of short duration (100 μs) presented independently to each ear through earphones with foam ear tips at a level of 90 dBnHL at a rate of 11.0/s. Filters were set at 100 and 3000 Hz. The resulting tracing,characterized by five waves, represents the activity of the auditory pathway between the most distal part of the auditory nerve and the inferior quadrigeminal tubercle. Finally, P300 auditory cortical potentials are measured as part of the auditory assessment. Using the same electrode montage as used in ABR, burst stimuli were used, 50-ms tones presented at a rate of 1.1/s to the right ear through an insert phone. For each condition, the standard stimulus was presented with 80% probability and the target stimuli was presented randomly with 20% probability.

## Current status of the study

We are collecting data for the initial feasibility study. Our current sample is mainly composed of women. Therefore, a key focus for the following RCT is to ensure that the sample groups are as balanced as possible, both in terms of group sizes and gender distribution. Achieving this balance is crucial to the validity of our findings, as it helps to minimize potential confounding variables that could arise from disproportionate representation of genders or uneven group sizes.

By reaching a well-balanced sample, we aim to strengthen the reliability of our comparisons between the experimental group (involved in music and other activities) and the control group (with sporadic non-musical activity or no activity). This approach is particularly important given the known variability in BDNF levels, which can be influenced by a range of factors including gender. The literature suggests that BDNF expression may differ between males and females, potentially due to hormonal influences; for a review, see Chan & Ye (2017). Therefore, by carefully balancing the gender percentages within each group, we aim to control for these potential differences and ensure that any observed effects can be more confidently attributed to the intervention itself, rather than to underlying gender-related variations.

We will also closely monitor the distribution of other demographic factors, as well as health conditions and medications regularly consumed, to further ensure that the groups are comparable. This meticulous approach to sampling is intended to enhance the robustness of our study, allowing us to draw more accurate and generalizable conclusions about the impact of the intervention on BDNF levels.

## Analysis

### 1. Analysis of BDNF levels

As validated commercial ELISA kits to measure BDNF in saliva samples are currently lacking, several studies have explored the possibility to extend the use of commercially available BDNF ELISA kits validated for plasma and serum, for the quantification of BDNF in saliva. Unfortunately, the significantly reduced levels of salivary versus circulating levels of BDNF, have resulted in the inability of the abovementioned kits to detect concentrations above the minimum detection threshold. In light of this, Mandel and colleagues (2011) have developed a sandwich ELISA procedure optimized for the measurement of BDNF in saliva. An analysis is currently underway to check whether, to date, commercial kits exist that have a detection range within which the expected salivary concentration of BDNF (median = 618 pg/ml) falls (**Figure 2**). If no such kits are identified, measurements will be performed with the ELISA optimized by Mandel (2011). Statistical analysis was performed using GraphPad Prism v8.4 software. The concentration of BDNF (pg/ml) in each sample will be calculated in accordance with a standard curve. The data will be expressed as means ± standard error of the mean (SEM).

**Figure 2.**
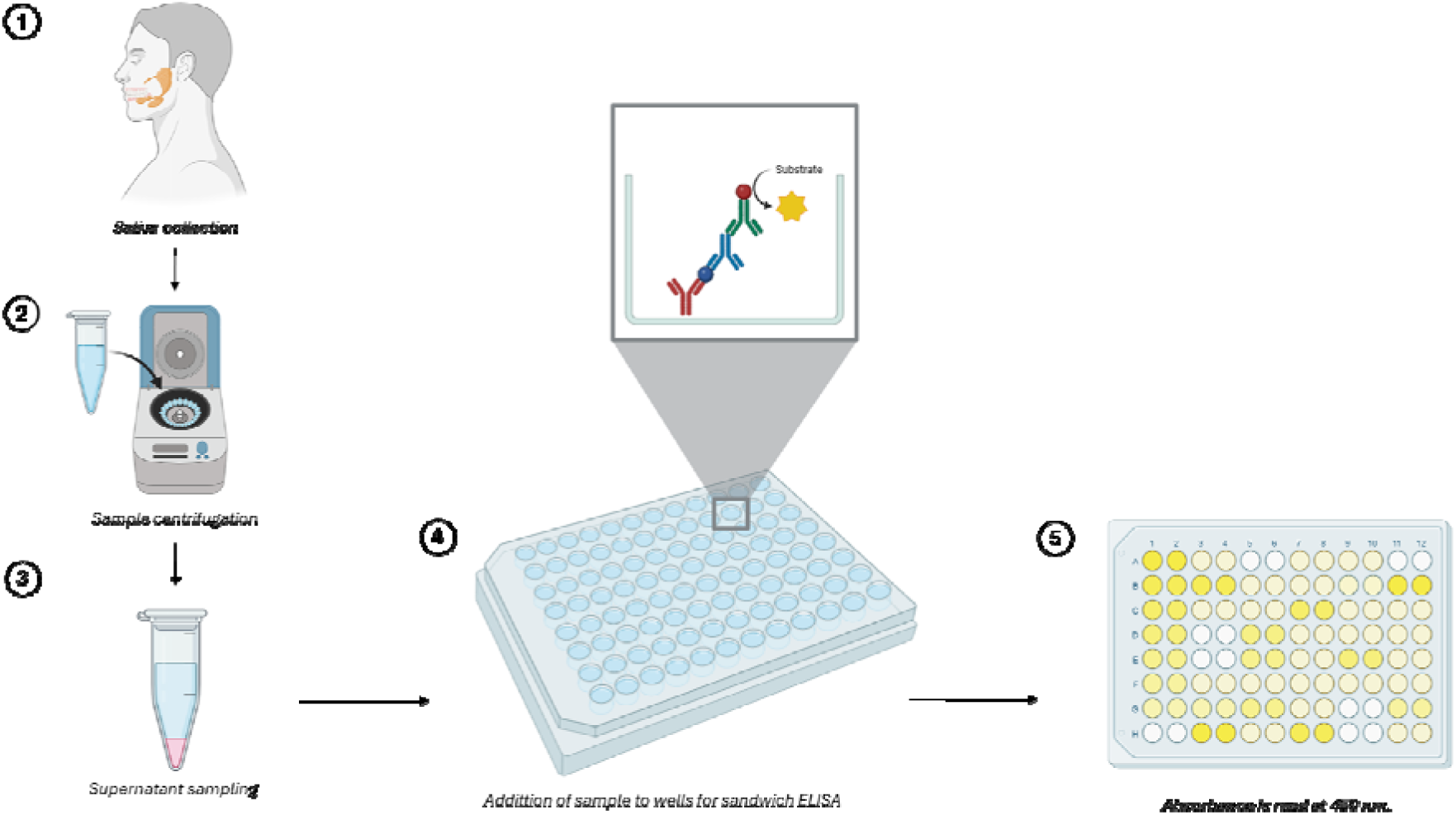
Schematic representation of the experimental protocol: (1-3) The saliva sample will be acquired, processed and stored as described (4) The sample will be added to a microtiter plate that has an antibody specific for BDNF immobilized on the bottom. The analyte present in the sample will bind to the antibody. A second antibody, conjugated to an enzyme, will bind to the analyte captured by the first antibody. Subsequently, a substrate for the enzyme will be added. The reaction between the enzyme and the substrate generates a colorimetric signal, which is proportional to the amount of analyte present in the sample. (5) The absorbance is then measured using a spectrophotometer, and the results are compared with a standard sample in order to quantify the analyte (Image was created with Biorender free version).

### 2. Statistical analysis

To evaluate differences in BDNF levels and other measurements between elderly individuals who participate regularly in group activities, such as choir and other activities, versus those who engage sporadically in non-musical activities, we will conduct comprehensive statistical analysis by means of Jamovi software as well as R Studio. Specifically, given the longitudinal design and the complexity of our study, we will use mixed-effects models to analyze the data. These models will account for both fixed effects, such as group membership, age, frailty score, and time, as well as random effects to capture individual variability. We will also include residential areas (urban vs. suburban) as a random effect to explore its potential influence on the outcomes. Mixed-effects models will be also used to examine how changes over time interact for each group with the different variables, including the effects of physical activity patterns, acoustic awareness, level of socio-psychological prosperity and multidimensional frailty. Mediation analyses will be also computed to deepen how the regular participation in group activities influence the outcomes of interest, thus supporting the direction of causal relationships. This comprehensive approach will allow us to see if and how all these variables are interrelated, and to assess the impact of regular versus sporadic activities, while also considering how perceptual levels and lifestyle may affect BDNF levels and related outcomes in relation to individual frailty conditions.

### 3. Power analysis

We calculated that a minimum of n = 26 participants per cohort was needed through an a priori sample size analysis using both G*power software (version 3.1.9.2, available online for free) and the *jpower* module from the Jamovi software (The jamovi project, 2023; *jamovi*. Version 2.4, retrieved from https://www.jamovi.org). We performed a t-test for ’Means: Difference between two independent means (two groups)’ to determine the required sample size based on α, power, and effect size. This analysis resulted in a total sample size of n = 52 participants, with 26 per group, based on the following parameters: two-tailed test, large effect size (d = 0.80), α error probability = 0.05, power (1 - β) = 0.80, and an equal allocation ratio between the two groups (N₂/N₁ = 1) (see **Table 5** and **Figure 3**).

**Figure 3.**
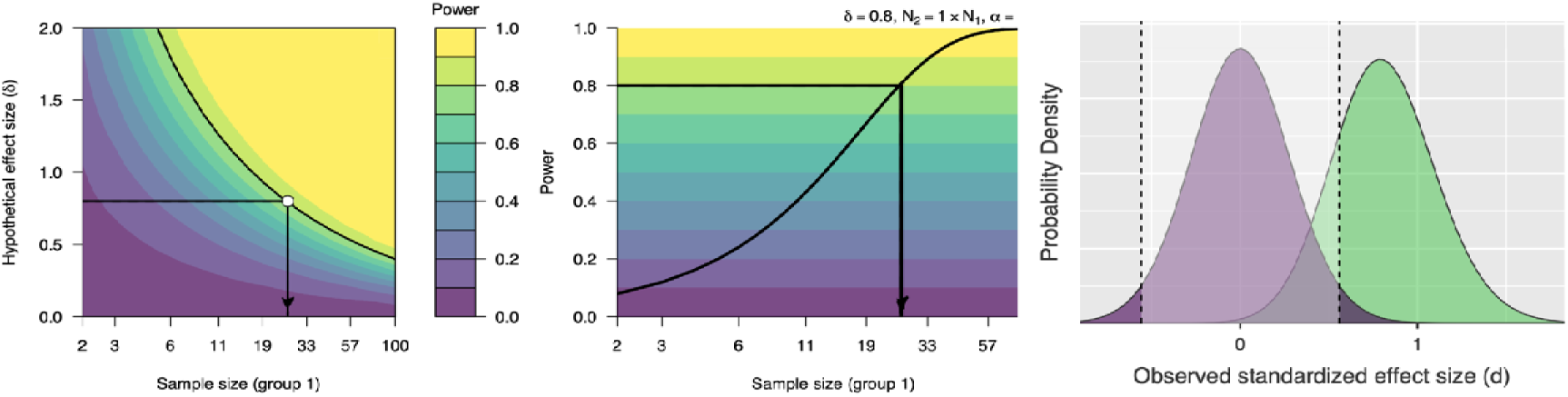
Power analysis

However, given the longitudinal nature of the study and the potential risk of dropouts, we plan to recruit 40 participants per group. Although the sample size might seem limited, previous studies on different types of physical training (i.e., aerobics, endurance, high-intensity interval training etc.) reporting significant effects in BDNF for intervention groups ranged from 7 to 47 participants (Seifert et al., 2010; Schmolesky et al., 2013; Yeh et al., 2015; Martínez-Díaz et al., 2020). Thus, the suggested sample seems adequate to identify main effects that are of relevant interest.

## Dissemination

Results of this research will be presented at national and international conferences focusing on geriatry or music research and published in a peer-reviewed journal.

## Data monitoring and management

A data monitoring committee is not considered as necessary because the study is of minimal risk. The behavioral and audiologic data from the digital questionnaires will be stored in an encrypted folder and the paper-and-pencil tests and questionnaires will be preserved in a locked cabinet and entered in an electronic file also stored in an encrypted folder. All saliva samples are kept on ice during the procedure and then centrifuged at 4000 rpm for 15 minutes at 4°C; the supernatant is then aliquoted and stored at -80°C until the assay is performed. The access to these data will be permitted to the research members only. Any adverse events will be reported by filling in the appropriate form. The data that support the findings of this study will be openly available on Zenodo repository.

## Expected outcomes

Based on our study design and previous literature, we expect to find significant results with regard to both the primary and secondary outcomes, with a special attention on BDNF being a measure under-investigated in the literature. Other primary and secondary outcomes related to cognition, auditory functions and multidimensional frailty are mentioned but will be treated in more detail in the future reports of this study.

The primary outcome on which we concentrate here is the BDNF level in the saliva. Specifically, we expect a difference in BDNF levels between the two groups (i.e., those who participate regularly in group activities, such as choir, and those who engage only sporadically or not at all), with stronger effects when including aspects of individual multidimensional frailty as a control variable. Previous research suggests that regular engagement in social and physically stimulating activities can lead to higher BDNF levels, particularly in elderly populations, where BDNF is associated with neuroplasticity and cognitive function. It should be noted that the referenced data pertains to BDNF levels in the circulation, rather than in saliva.

To date, no studies have been conducted that provide definitive evidence on the fluctuations of this protein within saliva over the course of an individual’s lifetime. Additionally, there is a lack of literature that suggests a correlation between salivary BDNF and circulating BDNF. We hypothesize that the group with regular participation will show a significant increase in BDNF levels over time compared to the sporadic activity group, reflecting the positive impact on brain health of sustained social and cognitive engagement. Also, we foresee the multidomain intervention as able in contrasting frailty aspects in aging or at least in keeping them stable over time, i.e., with no further deterioration in participants’ conditions.

Additional primary outcomes not related to BDNF focus on cognitive functions. First, based on previous research linking enriched environments to brain changes, we expect an improvement in general cognitive abilities in both active groups, assessed through behavioral tests. Second, we foresee an improvement for the choir group in near and far perceptual skills (assessed through musical, audiometric and auditory evoked potential tests) and hearing-related cognitive skills. In individuals of the intervention group, improvement is intended as decreased decline in the perceptuo-cognitive skills as opposed to controls. A concrete example of the expected changes in hearing-related cognitive skills is the speech-in-noise ability, which involves both basic auditory processing as well as higher-level cognitive functions. Indeed, choral singing seems to be particularly effective in improving speech perception in noise, especially through enhanced pitch discrimination, supporting the idea that even brief participation in a choir can be a valuable intervention to counter age-related hearing decline (Dubinsky et al., 2019).

By consequence, as secondary outcomes we expect to find significant correlation between BDNF levels and cognitive function, particularly in areas such as memory, fluid intelligence and attention. Considering the primary outcomes, we expect, of course, this relationship to be also linked to perceptual abilities. This analysis will be complemented with data coming from the neurophysiological assessment. For example, both latency and frequency of the P300 response, which reflects cognitive processes such as working memory and sensory discrimination, will be analyzed to assess the participants’ auditory processing abilities and cognitive function.

Moreover, we await a positive correlation between higher BDNF levels and physical activity, which is a well-known modulator of BDNF expression. Mediation analyses will show whether the increase of BDNF levels can mediate psychological well-being. In fact, we speculate that regular participation in group activities, especially those with a social component like choir, is likely to enhance socio-psychological well-being thanks to increased positive social interactions and stress reduction.

Further exploratory outcomes may involve the effects of control variables such as the area of residence, cognitive reserve and frailty level. We hypothesize urban environments might offer more opportunities for social interaction and engagement in activities, potentially leading to higher BDNF levels. The same applies to high levels of cognitive reserve and low levels of frailty, which we expect to facilitate a more active life in elderly people.

## Discussion

This ongoing study aims to evaluate the effects of a multidomain non-pharmacological intervention, including choral music training, on the BDNF levels, and on audiometric and neuropsychological measures in a sample of healthy older adults, assessed also for their multidimensional frailty. Changes in BDNF levels will then be correlated with a number of measures of cognitive, perceptual functioning and psychosocial well-being, to better understand the role of BDNF as a mediator in brain plasticity processes, especially in response to interventions aimed at boosting the cognitive and the social spheres. Moreover, the challenge of our project is to detect these changes in the elderly population, when neuroplasticity tends to be reduced, as a prevention strategy for multidimensional frailty.

In recent years, the idea that stimulating different domains affects brain plasticity and connectivity, especially in aging, has become increasingly evident (Nyberg et al., 2012; Bamidis et al., 2014; Lee et al., 2023). Indeed, some studies already showed that programmes combining both physical and cognitive activities can slow cognitive decline and reduce the risk of dementia more effectively than interventions focused on only one domain (e.g., Ngandu et al., 2015; Rosenberg et al., 2020). However, real studies investigating the effects of the multi-domain intervention on BDNF levels are currently scarce (with the exception, e.g., of Moon et al., 2022) and addressing this lack is one of the main objectives of this study.

Also, and more specifically, this study would fill the gap of the current absence of longitudinal studies investigating the effects of music training on BDNF. In fact, a number of studies have been conducted so far on the effects of physical activity on BDNF (e.g., Seifert et al., 2010; Schmolesky et al., 2013; Yeh et al., 2015; Martínez-Díaz et al., 2020) but there is no long-term evidence that music training can increase BDNF levels over time. However, it can be inferred as not only physical activity, but also repeated listening to music leads to higher levels of BDNF (Brattico et al., 2021).

Music training has unique characteristics that allow the simultaneous stimulation of several domains (i.e., perceptual-sensory, cognitive, motor, social, emotional etc.) (Herholz & Zatorre, 2012). Indeed, the reiterated practice of movements is accompanied by auditory feedback, which allows in turn to regulate the following movement (Zatorre et al., 2007). Interestingly, this also applies to choral training between the sound produced by the voice and the motor work of the larynx (Dichter et al., 2018). So that, similarly to sport, playing an instrument and singing activate the body, to the extent that they can provoke changes in the brain. For example, in music players bimanual coordination allows a better connection between the hemispheres (Jäncke et al., 2000; Burunat et al., 2015). This is why the motor domain is considered a ‘’near skill‘’ to music training (Bangert & Altenmüller, 2003) and why playing an instrument can be considered on a par with physical activity (Rodrigues et al., 2010; Wan & Schlaug, 2010). Then, it is reasonable to assume music training’s capacity to affect BDNF levels.

In addition to the physiological measures, in this study we are also carrying out perceptual, cognitive and audiologic assessments, as they are very interconnected domains and are also linked to the BDNF. Perceptual skills are crucial for keeping aware of one’s surroundings. Especially the loss of hearing, affecting two thirds of the elderly population, has been linked to the progressive reduction of brain volume and subsequent cognitive deterioration (Lin et al., 2011; Peelle et al., 2011; Eckert et al., 2012). Without proper perceptual functioning, stimuli from the environment cannot be processed correctly and this has inevitable, negative implications for the brain. Functional changes in the brain in response to auditory signals are also found to reduce hearing in noisy environments (Campbell & Sharma, 2013), a function associated with cognitive processes (e.g., executive functions) susceptible to neurodegeneration.

As music training is known to be an activity that benefits the auditory system and enhances abilities like auditory discrimination, attention and memorization (Kraus & Chandrasekaran, 2010; Besson et al., 2011; Moreno & Bidelman, 2014; Habibi et al., 2018; Alain et al., 2019) as well as speech-in-noise perception (Zendel et al., 2017; Dubinsky et al., 2019) it is able to keep perceptual and cognitive functions well trained. For all these reasons, we believe that the implementation of choral training, stimulating the memorisation of sounds, intonation seeking, involvement of the body and synchronization with others, both melodically and rhythmically, is a key addition in a non-pharmacological intervention. Although the far transfer of music training is still a debated topic (see, e.g., Sala & Gobet 2017; Schellenberg & Lima, 2023), it is fair to assume that the multimodality of music training can be a resource for the strengthening of a variety of skills.

Thanks to its full cognitive, motor and sensory involvement, we think choir training to be able to contribute to neuroplasticity and go beyond musical abilities, reducing the risk of several impairments and diseases in aging. Maintaining a good level of neuroplasticity, in turn, makes one capable of being adaptable and keep learning. Also, the social dimension of choir should not be underestimated. While many studies on multidomain interventions focus on home-based methods and neglect the social aspect (e.g., Belleville e al., 2020; Moon et al., 2022), choral training offers a cohesive group experience. This, as seen in previous studies (e.g., Pentikäinen et al., 2021; 2023), leads to an increase in social well-being. Such evidence, together with the strong relationship existing between BDNF and stress (see, e.g., Notaras & van den Buuse, 2020), makes it conceivable that engaging in music may lead to well-being through the increase of BDNF levels, which positive feeds back into psychosocial outcomes. We infer indeed that participating in choral training can give an all-round sense of fulfillment, despite the hardships of old age.

In our study, we set up an observational period of 9 months as a fair period to detect differences between the groups, based on previous literature. Previous studies, in fact, were able to show changes in BDNF levels after a similar amount of time, even if as a result of a different type of training (Seifert et al., 2010; Yeh et al., 2015; Moon et al., 2022). For music training, some studies showed improvements in older adults after 4-6 months, in cognitive functions (Bugos et al., 2007), as well as mood and quality of life (Seinfeld et al., 2013). Also, a few studies already showed that elderly undergoing more intensive music training show improvements in cognitive abilities (Guo et al., 2020; Okely et al., 2023), but more research is needed on this particular topic.

The procedures for analyzing salivary BDNF levels are subject to several limitations as well. The most significant challenge is the absence of commercial kits, which are not yet sufficiently sensitive for this detection. Furthermore, although saliva is an easily collected sample, it is a highly complex matrix that is influenced by numerous factors, including the circadian rhythm of secretion, diet and medication. In order to minimize these variables, we ensured that the sample was collected at the same daily time range (14:00 p.m. to 17:00 p.m.) and provided careful instructions to the participants on the behavior to adopt prior to sample collection.

In sum, we propose a study protocol for investigating the effects of a multi-domain intervention based on choir practice on the older population which implements a robust methodology and a longitudinal design by meticulously taking into account the individual intensity of the multidimensional activities conducted by the participants.

## Data Availability

All data produced in the present study are available upon reasonable request to the authors

## Acknowledgments

This research was supported by EU funding within the MUR PNRR A novel public-private alliance to generate socioeconomic, biomedical and technological solutions for an inclusive Italian ageing society (project n. PE00000015, AGE-IT).

## References

Adachi, N., Numakawa, T., Richards, M., Nakajima, S., & Kunugi, H. (2014). New insight in expression, transport, and secretion of brain-derived neurotrophic factor: Implications in brain-related diseases. World journal of biological chemistry, 5(4), 409.

Alain, C., Moussard, A., Singer, J., Lee, Y., Bidelman, G. M., & Moreno, S. (2019). Music and visual art training modulate brain activity in older adults. Frontiers in Neuroscience, 13, 182.

Armstrong, T., & Bull, F. (2006). Development of the world health organization global physical activity questionnaire (GPAQ). Journal of Public Health, 14, 66–70.

Angelucci, F., Ricci, V., Gelfo, F., Martinotti, G., Brunetti, M., Sepede, G., … & Caltagirone, C. (2014). BDNF serum levels in subjects developing or not post-traumatic stress disorder after trauma exposure. Brain and cognition, 84(1), 118–122.

Bamidis, P. D., Fissler, P., Papageorgiou, S. G., Zilidou, V., Konstantinidis, E. I., Billis, A. S., … & Kolassa, I. T. (2015). Gains in cognition through combined cognitive and physical training: the role of training dosage and severity of neurocognitive disorder. Frontiers in aging neuroscience, 7, 152.

Bangert, M., & Altenmüller, E. O. (2003). Mapping perception to action in piano practice: a longitudinal DC-EEG study. BMC neuroscience, 4, 1–14.

Barbabella, F., Cela, E., Socci, M., Lucantoni, D., Zannella, M., & Principi, A. (2022). Active ageing in Italy: A systematic review of national and regional policies. International journal of environmental research and public health, 19(1), 600.

Belleville, S., Cuesta, M., Bieler-Aeschlimann, M., Giacomino, K., Widmer, A., Mittaz Hager, A. G., … & Demonet, J. F. (2020). Rationale and protocol of the StayFitLonger study: a multicentre trial to measure efficacy and adherence of a home-based computerised multidomain intervention in healthy older adults. BMC geriatrics, 20, 1–17.

Bergman-Nutley, S., Darki, F., & Klingberg, T. (2014). Music practice is associated with development of working memory during childhood and adolescence. FRONTIERS IN HUMAN NEUROSCIENCE, 7.

Biasutti, M., & Mangiacotti, A. (2021). Music training improves depressed mood symptoms in elderly people: a randomized controlled trial. The International Journal of Aging and Human Development, 92(1), 115–133.

Bidelman, G. M., & Alain, C. (2015). Musical training orchestrates coordinated neuroplasticity in auditory brainstem and cortex to counteract age-related declines in categorical vowel perception. Journal of Neuroscience, 35(3), 1240–1249.

Bidelman, G. M., Weiss, M. W., Moreno, S., & Alain, C. (2014). Coordinated plasticity in brainstem and auditory cortex contributes to enhanced categorical speech perception in musicians. European Journal of Neuroscience, 40(4), 2662–2673.

Binder, J. C., Martin, M., Zöllig, J., Röcke, C., Mérillat, S., Eschen, A., … & Shing, Y. L. (2016). Multi-domain training enhances attentional control. Psychology and Aging, 31(4), 390.

Bonetti, L., Bruzzone, S. E. P., Paunio, T., Kantojarvi, K., Kliuchko, M., Vuust, P., Palva, S., & Brattico, E. (2023). Moderate associations between BDNF Val66Met gene polymorphism, musical expertise, and mismatch negativity. HELIYON, 9(5).

Boyke, J., Driemeyer, J., Gaser, C., Büchel, C., & May, A. (2008). Training-induced brain structure changes in the elderly. Journal of Neuroscience, 28(28), 7031–7035.

Brattico, E., Bonetti, L., Ferretti, G., Vuust, P., & Matrone, C. (2021). Putting cells in motion: Advantages of endogenous boosting of BDNF production. Cells, 10(1), 1–16.

Brigadski, T., & Leßmann, V. (2020). The physiology of regulated BDNF release. Cell and Tissue Research, 382(1), 15–45.

Bugos, J. A., Bidelman, G. M., Moreno, S., Shen, D., Lu, J., & Alain, C. (2022). Music and Visual Art Training Increase Auditory-Evoked Theta Oscillations in Older Adults. BRAIN SCIENCES, 12(10).

Bugos, J. A., Perlstein, W. M., McCrae, C. S., Brophy, T. S., & Bedenbaugh, P. H. (2007). Individualized piano instruction enhances executive functioning and working memory in older adults. Aging and mental health, 11(4), 464–471.

Burunat, I., Brattico, E., Puoliväli, T., Ristaniemi, T., Sams, M., & Toiviainen, P. (2015). Action in perception: prominent visuo-motor functional symmetry in musicians during music listening. PloS one, 10(9), e0138238.

Campbell, J., & Sharma, A. (2013). Compensatory changes in cortical resource allocation in adults with hearing loss. Frontiers in systems neuroscience, 7, 71.

Chaddock-Heyman, L., Loui, P., Weng, T. B., Weisshappel, R., McAuley, E., & Kramer, A. F. (2021). Musical training and brain volume in older adults. Brain sciences, 11(1), 50.

Chaldarov, G. N., Tonchev, A. B., & Aloe, L. (2009). NGF and BDNF: from nerves to adipose tissue, from neurokines to metabokines. Rivista di psichiatria, 44(2), 79–87.

Chan, Y. W. F., and Kosinski, M. (2015). ICAR Project Wiki. Evanston, IL: International Cognitive Ability Resource (ICAR).

Chikahisa, S., Sei, H., Morishima, M., Sano, A., Kitaoka, K., Nakaya, Y., & Morita, Y. (2006). Exposure to music in the perinatal period enhances learning performance and alters BDNF/TrkB signaling in mice as adults. Behavioural brain research, 169(2), 312–319.

Clegg, A., Young, J., Iliffe, S., Rikkert, M. O., & Rockwood, K. (2013). Frailty in elderly people. The lancet, 381(9868), 752–762.

Colucci-D’Amato, L., Speranza, L., & Volpicelli, F. (2020). Neurotrophic factor BDNF, physiological functions and therapeutic potential in depression, neurodegeneration and brain cancer. International journal of molecular sciences, 21(20), 7777.

Condon, D. M., and Revelle, W. (2014). The international cognitive ability resource: development and initial validation of a public-domain measure. Intelligence 43, 52–64.

Cotman, C.W., Berchtold, N.C. and Christie, L.A. (2007) Exercise Builds Brain Health: Key Roles of Growth Factor Cascades and Inflammation. Trends in Neurosciences, 30, 464–472.

Coulton, S., Clift, S., Skingley, A., & Rodriguez, J. (2015). Effectiveness and cost-effectiveness of community singing on mental health-related quality of life of older people: randomised controlled trial. The British Journal of Psychiatry, 207(3), 250–255.

Criscuolo, A., Pando-Naude, V., Bonetti, L., Vuust, P., & Brattico, E. (2022). An ALE meta-analytic review of musical expertise. Scientific reports, 12(1), 11726.

Degé, F., & Kerkovius, K. (2018). The effects of drumming on working memory in older adults. Annals of the New York Academy of Sciences, 1423(1), 242–250.

De Witte, M., Spruit, A., van Hooren, S., Moonen, X., & Stams, G. J. (2020). Effects of music interventions on stress-related outcomes: a systematic review and two meta-analyses. Health psychology review, 14(2), 294–324.

Diamond, M. C., Krech, D., & Rosenzweig, M. R. (1964). The effects of an enriched environment on the histology of the rat cerebral cortex. Journal of Comparative Neurology, 123(1), 111–119.

Dichter, B. K., Breshears, J. D., Leonard, M. K., & Chang, E. F. (2018). The control of vocal pitch in human laryngeal motor cortex. Cell, 174(1), 21–31.

Diener, E., Wirtz, D., Tov, W., Kim-Prieto, C., Choi, D. W., Oishi, S., & Biswas-Diener, R. (2010). New well-being measures: Short scales to assess flourishing and positive and negative feelings. Social indicators research, 97, 143–156.

Draganski, B., Gaser, C., Busch, V., Schuierer, G., Bogdahn, U., & May, A. (2004). Neuroplasticity: changes in grey matter induced by training. Nature, 427(6972), 311–312.

Driscoll, I., Martin, B., An, Y., Maudsley, S., Ferrucci, L., Mattson, M. P., & Resnick, S. M. (2012). Plasma BDNF is associated with age-related white matter atrophy but not with cognitive function in older, non-demented adults. PloS one, 7(4), e35217.

Dubinsky, E., Wood, E. A., Nespoli, G., & Russo, F. A. (2019). Short-Term Choir Singing Supports Speech-in-Noise Perception and Neural Pitch Strength in Older Adults With Age-Related Hearing Loss. Frontiers in neuroscience, 13, 1153.

Durany, N., Michel, T., Zöchling, R., Boissl, K. W., Cruz-Sánchez, F. F., Riederer, P., & Thome, J. (2001). Brain-derived neurotrophic factor and neurotrophin 3 in schizophrenic psychoses. Schizophrenia research, 52(1-2), 79–86.

Eckert, M. A., Cute, S. L., Vaden, K. I., Kuchinsky, S. E., & Dubno, J. R. (2012). Auditory cortex signs of age-related hearing loss. Journal of the Association for Research in Otolaryngology, 13, 703–713.

Edelmann, E., & Leßmann, V. (2018). Analyzing synaptic plasticity at the single cell level with STDP. Neuroforum, 24(3), A143–A150.

Egan, M. F., Kojima, M., Callicott, J. H., Goldberg, T. E., Kolachana, B. S., Bertolino, A., … & Weinberger, D. R. (2003). The BDNF val66met polymorphism affects activity-dependent secretion of BDNF and human memory and hippocampal function. Cell, 112(2), 257–269.

Elfving, B., Plougmann, P. H., & Wegener, G. (2010). Detection of brain-derived neurotrophic factor (BDNF) in rat blood and brain preparations using ELISA: pitfalls and solutions. Journal of neuroscience methods, 187(1), 73–77.

Ellis, R. J., Bruijn, B., Norton, A. C., Winner, E., & Schlaug, G. (2013). Training-mediated leftward asymmetries during music processing: a cross-sectional and longitudinal fMRI analysis. NeuroImage, 75, 97–107.

Erickson, K. I., Voss, M. W., Prakash, R. S., Basak, C., Szabo, A., Chaddock, L., … & Kramer, A. F. (2011). Exercise training increases size of hippocampus and improves memory. Proceedings of the national academy of sciences, 108(7), 3017–3022.

Foltran, R. B., & Diaz, S. L. (2016). BDNF isoforms: a round trip ticket between neurogenesis and serotonin?. Journal of neurochemistry, 138(2), 204–221.

Forti, L. N., Van Roie, E., Njemini, R., Coudyzer, W., Beyer, I., Delecluse, C., & Bautmans, I. (2015). Dose-and gender-specific effects of resistance training on circulating levels of brain derived neurotrophic factor (BDNF) in community-dwelling older adults. Experimental gerontology, 70, 144–149.

Gao, L., Zhang, Y., Sterling, K., & Song, W. (2022). Brain-derived neurotrophic factor in Alzheimer’s disease and its pharmaceutical potential. Translational neurodegeneration, 11(1), 4.

Gerstorf, D., Bertram, L., Lindenberger, U., Pawelec, G., Demuth, I., Steinhagen-Thiessen, E., & Wagner, G. G. (2016). Editorial. Gerontology, 62(3), 311–315.

Ghinelli, E., Johansson, J., Ríos, J. D., Chen, L. L., Zoukhri, D., Hodges, R. R., & Dartt, D. A. (2003). Presence and localization of neurotrophins and neurotrophin receptors in rat lacrimal gland. Investigative ophthalmology & visual science, 44(8), 3352–3357.

Glass, T. A., De Leon, C. F., Bassuk, S. S., & Berkman, L. F. (2006). Social engagement and depressive symptoms in late life: longitudinal findings. Journal of aging and health, 18(4), 604–628.

Hariri, A. R., Goldberg, T. E., Mattay, V. S., Kolachana, B. S., Callicott, J. H., Egan, M. F., & Weinberger, D. R. (2003). Brain-derived neurotrophic factor val66met polymorphism affects human memory-related hippocampal activity and predicts memory performance. Journal of Neuroscience, 23(17), 6690–6694.

Guastafierro, E., Rocco, I., Quintas, R., Corso, B., Minicuci, N., Vittadello, F., … & Sattin, D. (2022). Identification of determinants of healthy ageing in Italy: results from the national survey IDAGIT. Ageing & Society, 42(8), 1760–1780.

Guo, X., Yamashita, M., Suzuki, M., Ohsawa, C., Asano, K., Abe, N., … & Sekiyama, K. (2021). Musical instrument training program improves verbal memory and neural efficiency in novice older adults. Human Brain Mapping, 42(5), 1359–1375.

Habibi, A., Damasio, A., Ilari, B., Veiga, R., Joshi, A. A., Leahy, R. M., … & Damasio, H. (2018). Childhood music training induces change in micro and macroscopic brain structure: results from a longitudinal study. Cerebral Cortex, 28(12), 4336–4347.

Harrison, P., Collins, T., and Müllensiefen, D. (2017). Applying modern psychometric techniques to melodic discrimination testing: item response theory, computerised adaptive testing, and automatic item generation. Sci. Rep. 7, 1–18. doi: 10.1038/s41598-017-03586-z

Herholz, S. C., & Zatorre, R. J. (2012). Musical training as a framework for brain plasticity: behavior, function, and structure. Neuron, 76(3), 486–502.

Huang, T. Y., Chou, M. Y., Liang, C. K., Lin, Y. T., Chen, R. Y., & Wu, P. F. (2023). Physical activity plays a crucial role in multidomain intervention for frailty prevention. Aging Clinical and Experimental Research, 35(6), 1283–1292.

Hyde, K. L., Lerch, J., Norton, A., Forgeard, M., Winner, E., Evans, A. C., & Schlaug, G. (2009). Musical training shapes structural brain development. Journal of Neuroscience, 29(10), 3019–3025.

Ikenouchi, A., Okamoto, N., Hamada, S., Chibaatar, E., Fujii, R., Konishi, Y., … & Yoshimura, R. (2023). Association between salivary mature brain-derived neurotrophic factor and psychological distress in healthcare workers. Brain and Behavior, 13(12), e3278.

Italian National Institute of Health. (2023). Frailty in older adults. Retrieved from https://www.iss.it/

Jabusch, H. C., Alpers, H., Kopiez, R., Vauth, H., & Altenmüller, E. (2009). The influence of practice on the development of motor skills in pianists: A longitudinal study in a selected motor task. Human movement science, 28(1), 74–84.

Jäncke, L., Shah, N. J., & Peters, M. (2000). Cortical activations in primary and secondary motor areas for complex bimanual movements in professional pianists. Cognitive Brain Research, 10(1-2), 177–183.

Jasim, H., Carlsson, A., Hedenberg-Magnusson, B., Ghafouri, B., & Ernberg, M. (2018). Saliva as a medium to detect and measure biomarkers related to pain. Scientific reports, 8(1), 3220.

Johnson, J. K., Louhivuori, J., Stewart, A. L., Tolvanen, A., Ross, L., & Era, P. (2013). Quality of life (QOL) of older adult community choral singers in Finland. International psychogeriatrics, 25(7), 1055–1064.

Kagami, H., Hiramatsu, Y., Hishida, S., Okazaki, Y., Horie, K., Oda, Y., & Ueda, M. (2000). Salivary growth factors in health and disease. Advances in dental research, 14(1), 99–102.

Karege, F., Bondolfi, G., Gervasoni, N., Schwald, M., Aubry, J. M., & Bertschy, G. (2005). Low brain-derived neurotrophic factor (BDNF) levels in serum of depressed patients probably results from lowered platelet BDNF release unrelated to platelet reactivity. Biological psychiatry, 57(9), 1068–1072.

Karpova, N. N. (2014). Role of BDNF epigenetics in activity-dependent neuronal plasticity. Neuropharmacology, 76, 709–718.

Kojima, M., & Mizui, T. (2017). BDNF propeptide: a novel modulator of synaptic plasticity. Vitamins and Hormones, 104, 19–28.

Komulainen, Pirjo, Maria Pedersen, Tuomo Hänninen, Helle Bruunsgaard, Timo A. Lakka, Miia Kivipelto, Maija Hassinen, Tuomas H. Rauramaa, Bente K. Pedersen, and Rainer Rauramaa. (2008). "BDNF is a novel marker of cognitive function in ageing women: the DR’s EXTRA Study." Neurobiology of learning and memory 90, 596–603.

Kowiański, P., Lietzau, G., Czuba, E., Waśkow, M., Steliga, A., & Moryś, J. (2018). BDNF: a key factor with multipotent impact on brain signaling and synaptic plasticity. Cellular and molecular neurobiology, 38, 579–593.

Kraus, N., & Chandrasekaran, B. (2010). Music training for the development of auditory skills. Nature reviews neuroscience, 11(8), 599–605.

Lappe, C., Herholz, S. C., Trainor, L. J., & Pantev, C. (2008). Cortical plasticity induced by short-term unimodal and multimodal musical training. Journal of Neuroscience, 28(39), 9632–9639.

Larrouy-Maestri, P., Harrison, P., and Müllensiefen, D. (2019). The mistuning perception test: a new measurement instrument. Behav. Res. Methods 51, 663–675. doi: 10.3758/s13428-019-01225-1

Laske, C., Banschbach, S., Stransky, E., Bosch, S., Straten, G., Machann, J., … & Eschweiler, G. W. (2010). Exercise-induced normalization of decreased BDNF serum concentration in elderly women with remitted major depression. International Journal of Neuropsychopharmacology, 13(5), 595–602.

Lee, S., Harada, K., Bae, S., Harada, K., Makino, K., Anan, Y., … & Shimada, H. (2023). A non-pharmacological multidomain intervention of dual-task exercise and social activity affects the cognitive function in community-dwelling older adults with mild to moderate cognitive decline: A randomized controlled trial. Frontiers in aging neuroscience, 15, 1005410.

Lin, F. R., Ferrucci, L., Metter, E. J., An, Y., Zonderman, A. B., & Resnick, S. M. (2011). Hearing loss and cognition in the Baltimore Longitudinal Study of Aging. Neuropsychology, 25(6), 763–770.

Lippolis, M., Carlomagno, F., Campo, F. F., & Brattico, E. (2023). The Use of Music and Brain Stimulation in Clinical Settings: Frontiers and Novel Approaches for Rehabilitation in Pathological Aging. In The Theory and Practice of Group Therapy. IntechOpen.

Lipsky, R. H., & Marini, A. M. (2007). Brain-derived neurotrophic factor in neuronal survival and behavior-related plasticity. Annals of the New York Academy of Sciences, 1122(1), 130–143.

Liu, Q. R., Walther, D., Drgon, T., Polesskaya, O., Lesnick, T. G., Strain, K. J., … & Uhl, G. R. (2005). Human brain derived neurotrophic factor (BDNF) genes, splicing patterns, and assessments of associations with substance abuse and Parkinson’s Disease. American Journal of Medical Genetics Part B: Neuropsychiatric Genetics, 134(1), 93–103.

Lommatzsch, M., Zingler, D., Schuhbaeck, K., Schloetcke, K., Zingler, C., Schuff-Werner, P., & Virchow, J. C. (2005). The impact of age, weight and gender on BDNF levels in human platelets and plasma. Neurobiology of aging, 26(1), 115–123.

Lövdén, M., Bodammer, N. C., Kühn, S., Kaufmann, J., Schütze, H., Tempelmann, C., … & Lindenberger, U. (2010). Experience-dependent plasticity of white-matter microstructure extends into old age. Neuropsychologia, 48(13), 3878–3883.

Loui, P., Raine, L. B., Chaddock-Heyman, L., Kramer, A. F., & Hillman, C. H. (2019). Musical instrument practice predicts white matter microstructure and cognitive abilities in childhood. Frontiers in psychology, 10, 1198.

Lu, J., Moussard, A., Guo, S., Lee, Y., Bidelman, G. M., Moreno, S., Skrotzki, C., Bugos, J., Shen, D., Yao, D., & Alain, C. (2022). Music training modulates theta brain oscillations associated with response suppression. ANNALS OF THE NEW YORK ACADEMY OF SCIENCES, 1516(1), 212–221.

MacAulay, R. K., Edelman, P., Boeve, A., Sprangers, N., & Halpin, A. (2019). Group music training as a multimodal cognitive intervention for older adults. Psychomusicology: Music, Mind, and Brain, 29(4), 180.

MacGregor, C., Andrade, P. E., Forth, J., Frieler, K., and Müllensiefen, D. (2022). The rhythm ability test (RAT): A new test of rhythm memory in children and adults (Manuscript in preparation).

MacGregor, C., and Müllensiefen, D. (2019). The musical emotion discrimination task: a new measure for assessing the ability to discriminate emotions in music. Front. Psychol. 10:1955. doi: 10.3389/fpsyg.2019.01955

Mandel, A. L., Ozdener, H., & Utermohlen, V. (2009). Identification of pro-and mature brain-derived neurotrophic factor in human saliva. Archives of oral biology, 54(7), 689–695.

Mandel, A. L., Ozdener, H., & Utermohlen, V. (2011). Brain-derived neurotrophic factor in human saliva: ELISA optimization and biological correlates. Journal of Immunoassay and Immunochemistry, 32(1), 18–30.

Marie, D., Müller, C. A., Altenmüller, E., Van De Ville, D., Jünemann, K., Scholz, D. S., … & James, C. E. (2023). Music interventions in 132 healthy older adults enhance cerebellar grey matter and auditory working memory, despite general brain atrophy. Neuroimage: reports, 3(2), 100166.

Martínez-Díaz, I. C., Escobar-Muñoz, M. C., & Carrasco, L. (2020). Acute effects of high-intensity interval training on brain-derived neurotrophic factor, cortisol and working memory in physical education college students. International journal of environmental research and public health, 17(21), 8216.

Matthews, V. B., Åström, M. B., Chan, M. H. S., Bruce, C. R., Krabbe, K. S., Prelovsek, O., … & Febbraio, M. A. (2009). Brain-derived neurotrophic factor is produced by skeletal muscle cells in response to contraction and enhances fat oxidation via activation of AMP-activated protein kinase. Diabetologia, 52, 1409–1418.

Mattson, M. P., Duan, W., & Guo, Z. (2003). Meal size and frequency affect neuronal plasticity and vulnerability to disease: cellular and molecular mechanisms. Journal of neurochemistry, 84(3), 417–431.

McDonald, M. W., Hayward, K. S., Rosbergen, I. C., Jeffers, M. S., & Corbett, D. (2018). Is environmental enrichment ready for clinical application in human post-stroke rehabilitation?. Frontiers in behavioral neuroscience, 12, 135.

Meuchel, L. W., Thompson, M. A., Cassivi, S. D., Pabelick, C. M., & Prakash, Y. S. (2011). Neurotrophins induce nitric oxide generation in human pulmonary artery endothelial cells. Cardiovascular research, 91(4), 668–676.

Minutillo, A., Panza, G. & Mauri, M.C.(2021) Retraction Note: Musical practice and BDNF plasma levels as a potential marker of synaptic plasticity: an instrument of rehabilitative processes. Neurol Sci 42, 3067.

Miranda, M., Morici, J. F., Zanoni, M. B., & Bekinschtein, P. (2019). Brain-derived neurotrophic factor: a key molecule for memory in the healthy and the pathological brain. Frontiers in cellular neuroscience, 13, 472800.

Mizoguchi, H., Nakade, J., Tachibana, M., Ibi, D., Someya, E., Koike, H., … & Yamada, K. (2011). Matrix metalloproteinase-9 contributes to kindled seizure development in pentylenetetrazole-treated mice by converting pro-BDNF to mature BDNF in the hippocampus. Journal of Neuroscience, 31(36), 12963–12971.

Mizoguchi, Y., Yao, H., Imamura, Y., Hashimoto, M., & Monji, A. (2020). Lower brain-derived neurotrophic factor levels are associated with age-related memory impairment in community-dwelling older adults: the Sefuri study. Scientific Reports, 10(1), 16442.

Moon, S.Y., Kim, S., Choi, S.H. et al. (2022). Impact of Multidomain Lifestyle Intervention on Cerebral Cortical Thickness and Serum Brain-Derived Neurotrophic Factor: the SUPERBRAIN Exploratory Sub-study. Neurotherapeutics 19, 1514–1525

Mora F. (2013). Successful brain aging: plasticity, environmental enrichment, and lifestyle. Dialogues in clinical neuroscience, 15(1), 45–52.

Moreno, S., & Bidelman, G. M. (2014). Examining neural plasticity and cognitive benefit through the unique lens of musical training. Hearing research, 308, 84–97.

Morrison, J. H., & Baxter, M. G. (2012). The ageing cortical synapse: hallmarks and implications for cognitive decline. Nature Reviews Neuroscience, 13(4), 240–250.

Murukesu, R. R., Singh, D. K. A., Shahar, S., & Subramaniam, P. (2020). A multi-domain intervention protocol for the potential reversal of cognitive frailty:“WE-RISE” randomized controlled trial. Frontiers in public health, 8, 471.

Nair, P. S., Raijas, P., Ahvenainen, M., Philips, A. K., Ukkola-Vuoti, L., & Järvelä, I. (2021). Music-listening regulates human microRNA expression. Epigenetics, 16(5), 554–566.

Nasreddine, Z. S., Phillips, N. A., Bédirian, V., Charbonneau, S., Whitehead, V., Collin, I., … & Chertkow, H. (2005). The Montreal Cognitive Assessment, MoCA: a brief screening tool for mild cognitive impairment. Journal of the American Geriatrics Society, 53(4), 695–699.

Nazam, F., Shaikh, S., Nazam, N., Alshahrani, A. S., Hasan, G. M., & Hassan, M. I. (2021). Mechanistic insights into the pathogenesis of neurodegenerative diseases: towards the development of effective therapy. Molecular and Cellular Biochemistry, 476, 2739–2752.

Nettiksimmons, J., Simonsick, E. M., Harris, T., Satterfield, S., Rosano, C., Yaffe, K., & Health ABC Study. (2014). The associations between serum brain-derived neurotrophic factor, potential confounders, and cognitive decline: a longitudinal study. PLoS One, 9(3), e91339.

Ngandu, T., Lehtisalo, J., Solomon, A., Levälahti, E., Ahtiluoto, S., Antikainen, R., … & Kivipelto, M. (2015). A 2-year multidomain intervention of diet, exercise, cognitive training, and vascular risk monitoring versus control to prevent cognitive decline in at-risk elderly people (FINGER): A randomised controlled trial. The Lancet, 385(9984), 2255–2263.

Notaras, M., & van den Buuse, M. (2020). Neurobiology of BDNF in fear memory, sensitivity to stress, and stress-related disorders. Molecular psychiatry, 25(10), 2251–2274.

Nucci, M., Mapelli, D., & Mondini, S. (2012). Cognitive Reserve Index questionnaire (CRIq): a new instrument for measuring cognitive reserve. Aging clinical and experimental research, 24(3), 218–226.

Nyberg, L., Lövdén, M., Riklund, K., Lindenberger, U., & Bäckman, L. (2012). Memory aging and brain maintenance. Trends in cognitive sciences, 16(5), 292–305.

Okely, J. A., Cox, S. R., Deary, I. J., Luciano, M., & Overy, K. (2023). Cognitive aging and experience of playing a musical instrument. Psychology and Aging.

Olszewska, A. M., Gaca, M., Herman, A. M., Jednoróg, K., & Marchewka, A. (2021). How musical training shapes the adult brain: Predispositions and neuroplasticity. Frontiers in Neuroscience, 15, 630829.

Palasz, E., Wysocka, A., Gasiorowska, A., Chalimoniuk, M., Niewiadomski, W., & Niewiadomska, G. (2020). BDNF as a promising therapeutic agent in Parkinson’s disease. International journal of molecular sciences, 21(3), 1170.

Palomino, A., Vallejo-Illarramendi, A., González-Pinto, A., Aldama, A., González-Gómez, C., Mosquera, F., … & Matute, C. (2006). Decreased levels of plasma BDNF in first-episode schizophrenia and bipolar disorder patients. Schizophrenia research, 86(1-3), 321–322.

Pandit, P., Crewther, B., Cook, C., Punyadeera, C., & Pandey, A. K. (2024). Sensing methods for stress biomarker detection in human saliva: a new frontier for wearable electronics and biosensing. Materials Advances.

Park, D. C., & Reuter-Lorenz, P. (2009). The adaptive brain: aging and neurocognitive scaffolding. Annual review of psychology, 60(1), 173–196.

Peelle, J. E., Troiani, V., Grossman, M., & Wingfield, A. (2011). Hearing loss in older adults affects neural systems supporting speech comprehension. Journal of neuroscience, 31(35), 12638–12643.

Peng, S., Wuu, J., Mufson, E. J., & Fahnestock, M. (2005). Precursor form of brain-derived neurotrophic factor and mature brain-derived neurotrophic factor are decreased in the pre-clinical stages of Alzheimer’s disease. Journal of neurochemistry, 93(6), 1412–1421.

Pentikainen, E., Kimppa, L., Pitkaniemi, A., Lahti, O., & Sarkamo, T. (2023). Longitudinal effects of choir singing on aging cognition and wellbeing: a two-year follow-up study. FRONTIERS IN HUMAN NEUROSCIENCE, 17.

Pentikäinen, E., Pitkäniemi, A., Siponkoski, S. T., Jansson, M., Louhivuori, J., Johnson, J. K., … & Särkämö, T. (2021). Beneficial effects of choir singing on cognition and well-being of older adults: Evidence from a cross-sectional study. PloS one, 16(2), e0245666.

Perus, L., Busto, G. U., Mangin, J. F., Le Bars, E., & Gabelle, A. (2022). Effects of preventive interventions on neuroimaging biomarkers in subjects at-risk to develop Alzheimer’s disease: A systematic review. Frontiers in Aging Neuroscience, 14, 1014559.

Pickersgill, J. W., Turco, C. V., Ramdeo, K., Rehsi, R. S., Foglia, S. D., & Nelson, A. J. (2022). The Combined Influences of Exercise, Diet and Sleep on Neuroplasticity. Frontiers in psychology, 13, 831819.

Pilotto, A., Custodero, C., Maggi, S., Polidori, M. C., Veronese, N., & Ferrucci, L. (2020). A multidimensional approach to frailty in older people. Ageing research reviews, 60, 101047.

Pilotto, A., Veronese, N., Quispe Guerrero, K. L., Zora, S., Boone, A. L., Puntoni, M., … & EFFICHRONIC Consortium. (2019). Development and validation of a self-administered multidimensional prognostic index to predict negative health outcomes in community-dwelling persons. Rejuvenation Research, 22(4), 299–305.

Polacchini, A., Metelli, G., Francavilla, R., Baj, G., Florean, M., Mascaretti, L. G., & Tongiorgi, E. (2015). A method for reproducible measurements of serum BDNF: comparison of the performance of six commercial assays. Scientific reports, 5(1), 17989.

Poscia, A., Falvo, R., La Milia, D. I., Collamati, A., Pelliccia, F., Kowalska-Bobko, I., … & Moscato, U. (2017). Healthy ageing: happy ageing: health Promotion for Older People in Italy.

Reichardt, L. F. (2006). Neurotrophin-regulated signalling pathways. Philosophical Transactions of the Royal Society B: Biological Sciences, 361(1473), 1545–1564.

Reuter-Lorenz, P. A., & Park, D. C. (2014). How does it STAC up? Revisiting the scaffolding theory of aging and cognition. Neuropsychology review, 24, 355–370.

Rodrigues, A. C., Loureiro, M. A., & Caramelli, P. (2010). Musical training, neuroplasticity and cognition. Dementia & neuropsychologia, 4(4), 277–286.

Rosenberg, A., Mangialasche, F., Ngandu, T. et al. (2020). Multidomain Interventions to Prevent Cognitive Impairment, Alzheimer’s Disease, and Dementia: From FINGER to World-Wide FINGERS. J Prev Alzheimers Dis 7, 29–36.

Rosenberg, A., Ngandu, T., Rusanen, M., Antikainen, R., Bäckman, L., Havulinna, S., Hänninen, T., Laatikainen, T., Lehtisalo, J., Levälahti, E., Lindström, J., Paajanen, T., Peltonen, M., Soininen, H., Stigsdotter-Neely, A., Strandberg, T., Tuomilehto, J., Solomon, A., & Kivipelto, M. (2018). Multidomain lifestyle intervention benefits a large elderly population at risk for cognitive decline and dementia regardless of baseline characteristics: The FINGER trial. Alzheimer’s & dementia : the journal of the Alzheimer’s Association, 14(3), 263–270.

Rosenzweig, M. R., Krech, D., Bennett, E. L., & Diamond, M. C. (1962). Effects of environmental complexity and training on brain chemistry and anatomy: a replication and extension. Journal of comparative and physiological psychology, 55(4), 429.

Rowe, J. W., & Kahn, R. L. (1997). Successful aging. The Gerontologist, 37(4), 433–440.

Sala, G., & Gobet, F. (2017). When the music’s over. Does music skill transfer to children’s and young adolescents’ cognitive and academic skills? A meta-analysis. Educational Research Review, 20, 55–67.

Saruta, J., Lee, T., Shirasu, M., Takahashi, T., Sato, C., Sato, S., & Tsukinoki, K. (2010). Chronic stress affects the expression of brain-derived neurotrophic factor in rat salivary glands. Stress, 13(1), 53–60.

Saruta, J., Fujino, K., To, M., & Tsukinoki, K. (2012). Expression and localization of brain-derived neurotrophic factor (BDNF) mRNA and protein in human submandibular gland. Acta histochemica et cytochemica, 45(4), 211–218.

Scharfman, H. E., & MacLusky, N. J. (2005). Similarities between actions of estrogen and BDNF in the hippocampus: coincidence or clue?. Trends in neurosciences, 28(2), 79–85.

Schlaug, G. (2009). Music, musicians, and brain plasticity. Oxford handbook of music psychology, 197–207.

Schega, L., Peter, B., Brigadski, T., Leßmann, V., Isermann, B., Hamacher, D., & Törpel, A. (2016). Effect of intermittent normobaric hypoxia on aerobic capacity and cognitive function in older people. Journal of Science and Medicine in Sport, 19(11), 941–945.

Schellenberg, E. G., & Lima, C. F. (2024). Music training and nonmusical abilities. Annual Review of Psychology, 75(1), 87–128.

Schlaug, G. (2015). Musicians and music making as a model for the study of brain plasticity. Progress in brain research, 217, 37–55.

Schmolesky, M. T., Webb, D. L., & Hansen, R. A. (2013). The effects of aerobic exercise intensity and duration on levels of brain-derived neurotrophic factor in healthy men. Journal of sports science & medicine, 12(3), 502.

Schneider, P., Engelmann, D., Groß, C., Bernhofs, V., Hofmann, E., Christiner, M., … & Seither-Preisler, A. (2023). Neuroanatomical disposition, natural development, and training-induced plasticity of the human auditory system from childhood to adulthood: a 12-year study in musicians and nonmusicians. Journal of Neuroscience, 43(37), 6430–6446.

Schneider, N., & Yvon, C. (2013). A review of multidomain interventions to support healthy cognitive ageing. The Journal of nutrition, health and aging, 17(3), 252–257.

Seifert, T., Brassard, P., Wissenberg, M., Rasmussen, P., Nordby, P., Stallknecht, B., … & Secher, N. H. (2010). Endurance training enhances BDNF release from the human brain. American Journal of Physiology-Regulatory, Integrative and Comparative Physiology, 298(2), R372–R377.

Seinfeld, S., Figueroa, H., Ortiz-Gil, J., & Sanchez-Vives, M. V. (2013). Effects of music learning and piano practice on cognitive function, mood and quality of life in older adults. Frontiers in psychology, 4, 810.

Seither-Preisler, A., Parncutt, R., & Schneider, P. (2014). Size and synchronization of auditory cortex promotes musical, literacy, and attentional skills in children. Journal of Neuroscience, 34(33), 10937–10949.

Shimada, H., Makizako, H., Doi, T., Yoshida, D., Tsutsumimoto, K., Anan, Y., … & Suzuki, T. (2014). A large, cross-sectional observational study of serum BDNF, cognitive function, and mild cognitive impairment in the elderly. Frontiers in aging neuroscience, 6, 69.

Shimizu, E., Hashimoto, K., Okamura, N., Koike, K., Komatsu, N., Kumakiri, C., … & Iyo, M. (2003). Alterations of serum levels of brain-derived neurotrophic factor (BDNF) in depressed patients with or without antidepressants. Biological psychiatry, 54(1), 70–75.

Sihvonen, A. J., Särkämö, T., Leo, V., Tervaniemi, M., Altenmüller, E., & Soinila, S. (2017). Music-based interventions in neurological rehabilitation. The Lancet Neurology, 16(8), 648–660.

Soya, H., Nakamura, T., Deocaris, C. C., Kimpara, A., Iimura, M., Fujikawa, T., Chang, H., McEwen, B. S., & Nishijima, T. (2007). BDNF induction with mild exercise in the rat hippocampus. Biochemical and biophysical research communications, 358(4), 961–967.

Suri, D., Veenit, V., Sarkar, A., Thiagarajan, D., Kumar, A., Nestler, E. J., … & Vaidya, V. A. (2013). Early stress evokes age-dependent biphasic changes in hippocampal neurogenesis, BDNF expression, and cognition. Biological psychiatry, 73(7), 658–666.

Taki, Y., Kinomura, S., Sato, K., Goto, R., Kawashima, R., & Fukuda, H. (2011). A longitudinal study of gray matter volume decline with age and modifying factors. Neurobiology of aging, 32(5), 907–915.

The jamovi project (2023). jamovi. (Version 2.4) [Computer Software]. Retrieved from https://www.jamovi.org.

Urtamo, A., Jyväkorpi, S. K., & Strandberg, T. E. (2019). Definitions of successful ageing: a brief review of a multidimensional concept. Acta bio-medica : Atenei Parmensis, 90(2), 359–363.

Van Praag, H., Kempermann, G., & Gage, F. H. (2000). Neural consequences of enviromental enrichment. Nature Reviews Neuroscience, 1(3), 191–198.

Varvarigou, M., Creech, A., Hallam, S., & McQueen, H. (2012). Benefits experienced by older people in group music-making activities. Journal of Applied Arts & Health, 3(2), 183–198.

Veronese, N., Noale, M., Cella, A., Custodero, C., Smith, L., Barbagelata, M., Maggi, S., Barbagallo, M., Sabbà, C., Ferrucci, L., & Pilotto, A. (2022). Multidimensional frailty and quality of life: data from the English Longitudinal Study of Ageing. Quality of life research : an international journal of quality of life aspects of treatment, care and rehabilitation, 31(10), 2985–2993.

Vrijen, C., Schenk, H. M., Hartman, C. A., & Oldehinkel, A. J. (2017). Measuring BDNF in saliva using commercial ELISA: Results from a small pilot study. Psychiatry research, 254, 340–346.

Walsh, E. I., Smith, L., Northey, J., Rattray, B., & Cherbuin, N. (2020). Towards an understanding of the physical activity-BDNF-cognition triumvirate: A review of associations and dosage. Ageing research reviews, 60, 101044.

Wan, C. Y., & Schlaug, G. (2013). Brain plasticity induced by musical training.

Wang, X., Soshi, T., Yamashita, M., Kakihara, M., Tsutsumi, T., Iwasaki, S., & Sekiyama, K. (2023). Effects of a 10-week musical instrument training on cognitive function in healthy older adults: implications for desirable tests and period of training. Frontiers in Aging Neuroscience, 15, 1180259.

Wang, N., & Tian, B. (2021). Brainl.Zderived neurotrophic factor in autoimmune inflammatory diseases. Experimental and Therapeutic Medicine, 22(5), 1–7.

Webster, M. J., Herman, M. M., Kleinman, J. E., & Weickert, C. S. (2006). BDNF and trkB mRNA expression in the hippocampus and temporal cortex during the human lifespan. Gene Expression Patterns, 6(8), 941–951.

White-Schwoch, T., Carr, K. W., Anderson, S., Strait, D. L., & Kraus, N. (2013). Older adults benefit from music training early in life: biological evidence for long-term training-driven plasticity. Journal of Neuroscience, 33(45), 17667–17674.

World Health Assembly. (2022). Resolution WHA75.8. Strengthening clinical trials to provide high-quality evidence on health interventions and to improve research quality and coordination. In Seventy-fifth World Health Assembly, Geneva, 22–28 May 2022.

Xing, Y., Chen, W., Wang, Y., Jing, W., Gao, S., Guo, D., … & Yao, D. (2016). Music exposure improves spatial cognition by enhancing the BDNF level of dorsal hippocampal subregions in the developing rats. Brain research bulletin, 121, 131–137.

Yeh, S. H., Lin, L. W., Chuang, Y. K., Liu, C. L., Tsai, L. J., Tsuei, F. S., … & Yang, K. D. (2015). Effects of Music Aerobic Exercise on Depression and Brain-Derived Neurotrophic Factor Levels in Community Dwelling Women. BioMed research international, 2015(1), 135893.

Zanto, T. P., Johnson, V., Ostrand, A., & Gazzaley, A. (2022). How musical rhythm training improves short-term memory for faces. Proceedings of the National Academy of Sciences, 119(41), e2201655119.

Zatorre, R. J., Chen, J. L., & Penhune, V. B. (2007). When the brain plays music: auditory–motor interactions in music perception and production. Nature reviews neuroscience, 8(7), 547–558.

Zatorre, R. J., Fields, R. D., & Johansen-Berg, H. (2012). Plasticity in gray and white: neuroimaging changes in brain structure during learning. Nature neuroscience, 15(4), 528–536.

Zendel, B. R., West, G. L., Belleville, S., & Peretz, I. (2019). Musical training improves the ability to understand speech-in-noise in older adults. Neurobiology of aging, 81, 102–115.

Zeng, Y., Tan, M., Kohyama, J., Sneddon, M., Watson, J. B., Sun, Y. E., & Xie, C. W. (2011). Epigenetic enhancement of BDNF signaling rescues synaptic plasticity in aging.

Zhao, M., Chen, L., Yang, J., Han, D., Fang, D., Qiu, X., … & Pan, H. (2018). BDNF Val66Met polymorphism, life stress and depression: A meta-analysis of gene-environment interaction. Journal of affective disorders, 227, 226–235.

Zuk, J., Benjamin, C., Kenyon, A., & Gaab, N. (2014). Behavioral and neural correlates of executive functioning in musicians and non-musicians. PloS one, 9(6), e99868.

